# SARS-CoV-2 among migrants and forcibly displaced populations: a rapid systematic review

**DOI:** 10.1101/2020.12.14.20248152

**Authors:** Maren Hintermeier, Hande Gencer, Katja Kajikhina, Sven Rohleder, Claudia Santos-Hövener, Marie Tallarek, Jacob Spallek, Kayvan Bozorgmehr

**Affiliations:** Section for Health Equity Studies and Migration, Department of General Practice and Health Services Research, University Hospital Heidelberg, Im Neuenheimer Feld 130.3, 69120 Heidelberg, Germany; Leibniz-Institute for Prevention Research and Epidemiology – BIPS, Department Prevention and Evaluation, Unit Social Epidemiology, Achterstr. 30, 28359 Bremen, Germany; Robert Koch Institute, Unit 28 Social Determinants of Health, Department of Health monitoring and Epidemiology, General-Pape-Straße 62, 12101, Berlin, Germany; Department of Population Medicine and Health Services Research, School of Public Health, Bielefeld University, D- 33501 Bielefeld, Germany; Department of Public Health, Brandenburg University of Technology Cottbus-Senftenberg, Universitätsplatz 1, 01968 Senftenberg, Germany

**Author notes:** **Address for correspondence** Prof. Dr. med. Kayvan Bozorgmehr (MSc), Department of Population Medicine and Health Services Research, School of Public Health, Bielefeld University, Postfach 10 01 31, D- 33501 Bielefeld, Germany.

**Keywords:** migrants, forcibly displaced, SARS-CoV-2, COVID-19, refugees, asylum seekers

## Abstract

The economic and health consequences of the COVID-19 pandemic pose a particular threat to vulnerable groups, such as migrants, particularly forcibly displaced populations. The aim of this review is (i) to synthesise the evidence on risk of infection and transmission among migrants, refugees, asylum seekers and internally displaced populations, and (ii) the effect of lockdown measures on these populations. We searched MEDLINE and WOS, preprint servers, and pertinent websites between 1st December 2019 and 26th June 2020. The included studies showed a high heterogeneity in study design, population, outcome and quality. The incidence risk of SARS-CoV-2 varied from 0·12% to 2·08% in non-outbreak settings and from 5·64% to 21·15% in outbreak settings. Migrants showed a lower hospitalisation rate compared to non-migrants. Negative impacts on mental health due to lockdown measures were found across respective studies. However, findings show a tenuous and heterogeneous data situation, showing the need for more robust and comparative study designs.

## 1. Introduction

The COVID-19 pandemic poses economic and health threats to people worldwide, especially to migrants and forcibly displaced populations, such as refugees, asylum seekers and internally displaced persons (IDP).[1] Policy measures taken to mitigate the spread of the severe acute respiratory syndrome coronavirus type 2 (SARS-CoV-2) may exacerbate poor health conditions among these populations, or exacerbate conditions which create or add to pre-existing vulnerabilities. Especially low-wage labour migrants and forcibly displaced populations often live in crowded accommodations where they share rooms as well as cooking and sanitary facilities with a number of people outside their own household. Physical distancing and recommended hygiene measures are not feasible in many such contexts.[2] Considering that 78 to 85% of human-to-human transmissions take place in family clusters and up to 10% on household level, these conditions are likely to increase the risk of infection and rapid dissemination among migrants and displaced populations.[3] Precarious working conditions may add to these vulnerabilities, mainly in manual labour jobs e.g., agricultural, or domestic work, that do not allow for protective measures like self-isolation, home-office, or physical distancing.[2, 4] This often leads to the loss of livelihoods due to the policy and lockdown measures taken, as it happened to many migrant workers in India under the national lockdown in March 2020.[5]

While the COVID-19 pandemic poses a threat to migrants and displaced populations, evidence on risk of infection, progress of disease, and effective prevention strategies is still lacking. An emerging body of evidence showed racial and ethnic disparities, with higher SARS-CoV-2 incidence in ethnic minorities compared to white persons.[6, 7] However, there is only a small number of studies investigating these aspects in migrants and displaced populations.

This review aims to synthesise the empirical evidence on risk of infection, transmission, development of disease, and risk of severe course of disease among migrants, refugees, asylum seekers and IDPs. The secondary objective is to review the evidence on the effects of lockdown measures on their health, and effective policy strategies to avert risks and negative outcomes. We thereby seek to summarise valuable information to inform future research in this field.

## 2. Methods

### 2.1 Search strategy and selection criteria

We conceived a rapid systematic review (PROSPERO registration number CRD42020195633) based on recommendations by the Cochrane Rapid Reviews Methods Group.[8]

To maintain quality while utilising resources efficiently, two underlying decisions have been made to accelerate the review process: the number of databases to search was restricted to two and the language of publication to English and German only.

The search strategy was twofold: Using scientific databases on the one hand and pertinent websites on the other to identify further relevant articles and grey literature. We conducted a systematic search query in MEDLINE via PubMed and Web of Science Core Collection (WOS) and searched in the major preprint servers medRxiv and bioRxiv to cover yet unpublished research results in this fast and early time of the current pandemic. The search included the time between 1st December 2019 and 26th June 2020 using English search terms for the study population and SARS-CoV-2 as exposure (for details on the search query see Supplementary File: Appendix A). Additional publications were identified on the websites of the International Organisation for Migration (IOM), the European Public Health Association (EUPHA), the World Health Organisation (WHO) COVID research, and the website of the systematic and living map on COVID-19 evidence of the Norwegian Institute of Public Health (NIPH). The website search was conducted by two independent researchers between the 25th and 29th of June 2020. The screening process was done in two steps. First, two researchers independently screened titles and abstracts by applying pre-defined inclusion/exclusion criteria (see table 1).

**Table 1:** Inclusion/Exclusion criteria.

| Inclusion/Exclusion criteria |  |  |
| --- | --- | --- |
| Criteria | Inclusion | Exclusion |
| Type of Population* | Refugees, asylum seekers, internally displaced people, and migrants | Unclear populations or reference to migrants and refugees without stratified results |
| Type of studies | • Empirical quantitative studies (cross-sectional, case-control. Cohort, intervention studies, ecological studies, modelling studies, epidemiological outbreak reports)<br>• Mixed-methods studies where the quantitative part is distinguishable<br>• Empirical qualitative studies (interviews, | Case series, theoretical research work; Policy analysis without empirical data |
|  | focus groups, ethnography) analysing health outcomes or impact of lock-down measures and evaluation strategies that include refugees and migrants<br>Identified rapid or systematic reviews will only be screened for their included articles; meta-analyses will be included on their own. |  |
| **Type of articles** | Published peer-reviewed articles and preprints identified, as well as official reports from IOM or EUPHA websites. | Grey literature except for official reports on mentioned websites. |
| **Focus of study** | Studies reporting on SARS-CoV-2 epidemiology (infection, disease), course of disease (hospitalization, ICU) or outcome of disease (mortality)<br>Studies reporting effects of lock-down measures, also on non-Covid health outcomes<br>Studies reporting effects of policies/strategies specifically enacted or implemented for refugees and migrants | Studies with no focus on SARS-CoV-2 or no reference to health outcomes. |
| **Type of outcome measure** | Measures of risk (prevalence ratios, incidence risk, risk ratios, rate ratios, odds ratios, hazard ratios) of SARS-CoV-2 infection, disease or outcome<br>Any measures of association between refugee and migrant health and lock-down measures or policies/strategies specifically enacted/implemented for refugees and migrants | Crude cases without denominators, proportions without denominators |
| **Geographical area** | No studies will be excluded based on geography. |  |
| **Language of publication** | Only studies published in English or German will be included (but searches will be conducted only in English). | Studies in other languages than English or German. |
| **Date of Publication** | Studies published since December 2019 |  |

Legend: \*Definitions of included populations: see detailed protocol registered in PROSPERO[9]

**Table 2:** Overview of included studies.

| Author(s) | Date of Publication | Type of Publication | Type of Study | Context of Study/Setting | Country of Study | Socioeconomic Development of Country/Region | Population | Sample Size (migrant group) | Outcome measures | Main outcomes as reported |
| --- | --- | --- | --- | --- | --- | --- | --- | --- | --- | --- |
| M. Irvine, D. Coombs, J. Skarha, B. Del Pozo, J. Rich, F. Taxman and T. C. Green | 2020 | Peer-reviewed | Modelling study | U.S. Immigration and Customs Enforcement (ICE) facilities, where detained immigrants are not housed individually or in cells as is true in many prison settings (detainees have more contacts with one another and with staff, thereby contributing to faster spread of infection than in correctional facilities) | USA | HIC | Detainees in 111 ICE facilities | N= 42,435 | Rate of SARS-CoV-2 Transmission<br>SARS-CoV-2 cases and proportion of detainees infected<br>Impacts on regional hospitalintensive care unit (ICU) capacity | First analysis assessing the impact of COVID-19 on ICE detainees and the wider communities likely to care for detainees. Across the 111 ICE facilities examined in the study, and considering all three scenarios, the cumulative number of ICU admissions by day 90 for over half of facilities would exceed hospital capacity within a 10-mile radius. The impact on hospital capacity is lower if the radius was expanded to 50 miles, hovering around 8% of facilities exceeding ICU demand. These models do not take into account other concerns that would strain ICE's operational capacity at a time when staffing is likely to be a concern, such as the difficulty and risk to staff of repeatedly transporting and guarding detainees up to 50 miles distant from their facilities of origin. The timing and peak number of cases are driven by facility size, which also are important when considering preventive interventions. |
| S. Truelove, O. Abraham, C. Altare, S. A. Lauer, K. H. Grantz, A. S. Azman and P. Spiegel | 2020 | Peer-reviewed | Modelling study | The Kutupalong-Balukhali Expansion Site, Bangladesh | Bangladesh | LIC | Rohingya refugees from Myanmar | n= 600.000 | Rate of SARS-CoV-2 transmission across three scenarios<br>SARS-CoV-2 cases and proportion of population infected<br>hospitalizations, deaths | Using a stochastic disease transmission model, the study estimated the number of people infected, hospitalizations, and deaths across three transmission scenarios after a successful introduction of SARS-CoV-2 into the Kutupalong-Balukhali Expansion Site. The introduction of SARS-CoV-2 into the Kutupalong-Balukhali Expansion Site or any other large refugee or internally displaced persons (IDPs) camp or settlement is likely to have serious consequences and overwhelm existing health systems. Even when transmission rates were assumed to be similar to that of influenza (low scenario), the necessary hospitalization capacities far exceeded the available capacities for the refugees in the expansion site in most simulations |
| M. Hariri, H. Rihawi, S. Safadi, M. A. McGlasson and W. Obaid | 2020 | Pre print | Modelling study | Camps and tented settlements along the Turkey-Syria border | Northwest Syria | LIC | Internally displaced persons (IDPs) in camps and tented settlements | n= 1.200.000 | SARS-CoV-2 caseload with the classification of mild, severe and critical cases in three different scenarios<br>ICU, deaths | Using a WHO forecasting tool, this study has identified COVID-19 case load, according to severity, and potential health system needs in NW Syria, an area that has been subjected to severe levels of violence, displacement and health system disruption during the nine years of the Syrian conflict. The study models use three scenarios. The Camp-population Scenario describes a situation whereby unmitigated spread in crowded displacement camps will lead to total health system collapse within the first four weeks of an outbreak. Considering that such a scenario can occur concurrently with Scenario Two (or a worse All-population Scenario Three not presented here whereby doubling rates and clinical attack rates would be higher), the epidemic outcomes can be catastrophic |
| I. Bojorquez, C. Infante, I. Vieitez, S. Larrea and C. Santoro | 13 May 2020 | Pre-print | Cross-sectional study (analytical) | Migrants in transit and asylum seekers in Mexico | Mexico | Upper MIC | Migrants, defined as persons who are part of mixed migrant flows (economic migrants and asylum seekers), of non-Mexican nationality, who are in Mexico or transiting through the country | n=74 | Frequency of suspect and positive cases hospitalisation, pneumonia, intubation cumulative incidence of suspected cases per 100,000 in migrants and non-migrants | This article presents an exploration of the COVID-19 pandemic among migrants and asylum seekers in Mexico, based on the available epidemiological surveillance information. Despite the limitations of the data, it is possible to draw some general conclusions. The first noticeable result is that 0.27% of the suspect cases registered in the database corresponded to migrants originating from Central America, the Caribbean, Venezuela or an African country, a very high percentage if one considers that international immigrants in Mexico were only about 0.11% of the population in mid-2020 in Mexico. When it is taken into consideration that the majority of international immigrants in Mexico come from the United States, the difference is even more striking. The calculation of cases per 100,000 shows a similar picture, particularly in scenario 1. [...] This could be an underestimation of the actual number, as not all migrants aiming to cross are registered in the waiting lists that are the source of this data. |
| K. Kumar, A. Mehra, S. Sahoo, R. Nehra and S. Grover | 2020 | Letter to the editor | Cross sectional study (descriptive) | Chandigarh, a Union Territory, in North India | India | Low MIC | Migrant workers identified by the Government of India, who were living in the shelter house or government authorized buildings | n=98 | Depression (PHQ2) Anxiety (GAD-2) perceived stress (PSS-4) Psychiatric morbidity | The present study suggests that about three-fourth (73.5 %) of the participants screened positive for either depression or anxiety. All the migrants who screened positive for anxiety also screened positive for depression, suggesting high co-morbidity. Additionally, about one-fifth of the participants screened positive for only depression. Additionally, on the self-designed questionnaire, a significant proportion of participants reported a marked increase in negative emotions and feelings such as loneliness, tension, frustration, low mood, irritability, fear, fear of death, and social isolation |
| D. Koh | 2020 | Short report | Cross sectional study (descriptive time-series) | Dormitory housing of migrant workers in Singapore | Singapore | HIC | Migrant workers in Singapore | not explicitly stated but said that "approximately 200.000 migrant workers" live in these 43 dormitories, plus 95.000 that live in 1200 factory-converted dormitories and 20.000 construction workers living in quarters at their worksite (N= 200.000+95.000+20.000 = 315.000). | Incident SARS-CoV-2 cases | From early April 2020, a marked escalation of new cases of COVID-19 was observed among low-skilled migrant workers living in dormitories in Singapore. By 6 May 2020, these infected workers formed 87.9% of the 20 198 cases of cases of COVID-19 confirmed in Singapore. Unsatisfactory housing and social overcrowding in accommodation for low-skilled migrant workers need to be addressed before any pandemic occurs. If this is not done, epicentres of the disease can arise in these housing areas. |
| I. Motta, R. Centis, L. D'Ambrosio, J.-M. García-García, D. Goletti, G. Gualano, F. Lipani, F. Palmieri, A. Sánchez-Montalvá, E. Pontali, G. Sotgiu, A. Spanevello, C. Stochino, E. Taberner, M. Tadolini, M. van den Boom, S. Villa, D. Visca and G. B. Migliori | 14 May 2020 | Peer-reviewed | Cohort study (descriptive) | 26 centres in Belgium, Brazil, France, Italy, Russia, Singapore, Spain, and Switzerland | Belgium, Brazil, France, Italy, Russia, Singapore, Spain, and Switzerland | HIC | Migrants diagnosed with TB in Belgium, Brazil, France, Italy, Russia, Singapore, Spain, and Switzerland | n=43 | Mortality rate | First report of patients dying with TB and COVID-19, including 69 patients from the two largest cohorts of co-infected patients available so far. Although the case-fatality rate was rather high (overall 10.6%, but 14.3% in the first cohort) and still preliminary (it can increase over time within both cohorts), the results seem consistent with those observed in other cohorts of COVID-19 patients. In general, all patients (except one) were aged >65 years, and were affected by >2 comorbidities. In all cases COVID-19 contributed to worsen the prognosis of TB patients and/or to cause death. |
| Mendez-Dominguez, N; Alvarez-Baeza, A; Carrillo, G | 15 June 2020 | Peer-reviewed | Epidemiologic, cross sectional study | Sentinel surveillance for SARS-CoV-2 cases in Mexico | Mexico | Upper MIC | Migrants defined as the totality of national and international migratory movements projected for the year 2020 from registry estimates | No specific sample size for migrant population given | Incidence, lethality, hospitalization, and confirmation of SARS-CoV-2 cases | In the present study, we observed that confirmation and hospitalizations were both more frequent in the states where more clinics and hospitals are available. At the same time, a proportion of patients moved from their state of residence to seek medical attention, according to the case-by-case results. [...] From the beginning of the outbreak, migration was significantly associated with incidence ratios according to state-cluster analyses. [...] Elderly patients had lower odds of being hospitalized, but were likely to die, while interstate migrants had more propensity to fatal outcomes, yet were less likely to be laboratory-confirmed. Age group laboratory-testing, if not corrected, could result in biased assumptions of severity and lethality among young patients. |
| T. D. A. Ly, V.<br>T. Hoang, N.<br>Goumballa, M.<br>Louni, N.<br>Canard, T. L.<br>Dao, H.<br>Medkour, A.<br>Borg, K. Bardy,<br>V. Esteves-Vieira, V.<br>Filosa, B.<br>Davoust, O.<br>Mediannikov,<br>P.-E. Fournier,<br>D. Raoult and P.<br>Gautret | 14 May<br>2020 | Pre-print | Cross-sectional<br>study<br>(descriptive) | Different shelters and<br>accommodation<br>centres in Marseille,<br>France | France | HIC | Asylum seekers | n=77 | symptoms,<br>temperature, PCR<br>SARS-CoV-19<br>carriage | First study addressing SARS-CoV-2 carriage among different precarious populations including homeless adults but also children and other hard-to-reach populations during the COVID-19 outbreak in France. The strength of our study is its large population size, with a high (78.9%) acceptance rate toward testing, particularly among individuals living in precarious conditions (92.1%) suggesting that this population is concerned about the disease. We found an overall 7.0% SARS-CoV-2 positivity rate, with most infected individuals among homeless people and employees working in homeless facilities, while no cases were found in asylum-seekers and in other people also living in precarious conditions |
| Guijarro, C;<br>Perez-<br>Fernandez, E;<br>Gonzalez-<br>Pineiro, B;<br>Melendez, V;<br>Goyanes, MJ;<br>Renilla, ME;<br>Casas, ML;<br>Sastre, I;<br>Velasco, M | 27 May<br>2020 | Pre-print | Population<br>based cohort<br>analytic study | Hospital Universitario<br>Fundación<br>Alcorcón, Spain | Spain | HIC | Migrants, defined by<br>their nationality | n= 20419 | Incidence rate of<br>PCR-confirmed cases<br>Relative risk ratios of<br>PCR+ cases in<br>different migrant<br>groups | The other major finding of our work is an apparent higher risk for COVID-19 for individuals from Sub-Saharan, Caribbean or Latin-American origin. All of them have equal access to the virtually universal health coverage available for Spaniards or migrants from other areas of the world.<br>In summary, we report a selective increased risk for COVID-19 among certain migrant populations in Spain: Sub-Saharan, Caribbean and Latin-America that is not related to unequal access to health care. These groups may deserve a particular attention, particularly when our country, as well as others, is beginning a de-escalation of social distancing measures |
| M. H. Chew, F.<br>H. Koh, J. T.<br>Wu, S.<br>Ngaserin, A.<br>Ng, B. C. Ong<br>and V. J. Lee | 31 May,<br>2020 | Letter to<br>the Editor | Cohort<br>analytic study | Dormitories in<br>Singapore | Singapore | HIC | Migrant workers<br>residing in a large<br>dormitory in<br>Singapore | n=5977 | PPV, NPV of clinical<br>parameters<br>(Symptoms, clinical<br>signs, Oxygen<br>saturation, heart rate)<br>prevalence rate of<br>PCR confirmed cases | Whereas 21% of the cohort (of 5977 foreign workers) were found to be positive for COVID-19, the true prevalence is likely higher if asymptomatic individuals are also assessed, with incidence up to 36% in other closed environments.<br>The strategy for containment in closed-living environments therefore would be to isolate symptomatic individuals, and to establish public health measures for social distancing |
| P. Rzymiski and M. Nowicki | 26. Apr 2020 | Peer-reviewed | Online cross-sectional study (descriptive) | Poznan University of Medical Sciences in Poland | Poland | HIC | Asian students at Poznan University of Medical Sciences in Poland | n=85 | Preconceptions related to SARS-CoV-2 | In conclusion, the reactions presented in this study clearly show that Asian students in regions yet unaffected by SARS-CoV-2 could have already experienced an uncomfortable level of prejudice in the public spaces encompassing transport, gastronomy, shopping, health services and university. Such behaviors can particularly affect those individuals of Asian origin who are tending to wear face masks. Overall, these findings underscore the responsibility of different parties in overcoming and preventing discrimination during outbreaks of infectious diseases. Firstly, universities that host Asian students and staff from abroad should support their students during the outbreak (and any other future epidemic emerging from Asian region) and protect them from harmful misconceptions. |
| Fakhar-e-Alam Kulyar, M; Bhutta, Zeeshan A; Shabbir, S; Akhtar, M | 25 April 2020 | Letter to the editor | Cross-sectional study (descriptive) | International students living in Hubei province | China | Upper MIC | International students living in Hubei province | n= 504 | Socio-psychological impact of SARS-CoV-2 on international students' daily life | This study revealed some specific socio-psychological experiences of respondents. However, it is also admirable that many of the international students were afraid during pandemic. This may be due to the fact that the respondents in affected areas paid more attention to the safety of their families [4]. Secondly, students with longer stay in China reported more concerns and consequences than the students who stayed for a short period of time. This may be associated with the respondent's age and their marital status |
| Lopez-Pena P, Austin Davis C, Mushfiq Mobarak A & Raihan S | 11 May 2020 | Pre-print | Cross-sectional study (population-based household survey) | Households across Cox's Bazar in Bangladeshi and Rohingya Refugees living in camps in Cox's Bazar | Bangladesh | LIC | Rohingya refugees | n=367 | Prevalence of SARS-CoV-2 Symptoms treatment seeking behaviour knowledge about SARS-CoV-2 and health behaviours | Camp residents report COVID-19 symptoms almost twice as frequently as members of the host community. We also document differences in self-reported non- COVID-19 symptoms, but these are not statistically significant. While this suggests that COVID-19 is much more prevalent in the refugee population, we cannot definitively exclude two alternative explanations. The first is that refugees experience higher rates of other common illnesses with overlapping symptoms. The second is that some refugees over report adverse life events and health outcomes, as some anecdotal evidence suggests. |
| Qiu, J., Shen, B., Zhao, M., Wang, Z., Xie, B., & Xu, Y | 29 February 2020 | Editorial | Nationwide large-scale survey (analytical) | Psychological distress in the general population of China during the tumultuous time of the COVID-19 epidemic | China | Upper MIC | Migrant workers | not stated | Psychological distress (only outcome for migrants) | Findings of this study suggest the following recommendations for future interventions: (1) more attention needs to be paid to vulnerable groups such as the young, the elderly, women and migrant workers; (2) accessibility to medical resources and the public health service system should be further strengthened and improved, particularly after reviewing the initial coping and management of the COVID-19 epidemic; (3) nationwide strategic planning and coordination for psychological first aid during major disasters, potentially delivered through telemedicine, should be established and (4) a comprehensive crisis prevention and intervention system including epidemiological monitoring, screening, referral and targeted intervention should be built to reduce psychological distress and prevent further mental health problems. |

Conflicts were resolved through discussion or, where no agreement was reached, by a third reviewer. In the next step, full text screening for eligible records was conducted by following the same procedure as the title and abstract screening. Due to the novelty of the current pandemic and the conception of this project as a rapid review, references of eligible articles were screened, and, alongside journal articles, peer-reviewed comments or letters to the editors were included, if they reported empirical data.

### 2.2 Quality assessment

The quality appraisal of included studies was carried out independently and in duplicate. Quantitative studies were assessed using the tool of the Effective Public Health Practice Project (EPHPP). Modelling studies were appraised using a self-developed instrument derived from existing tools.[10-13] The Covidence software was used for the screening of titles, abstract, and full-texts as well as for quality appraisal.

### 2.3 Data extraction and Outcomes

Data extraction was performed in Excel 2016 by one reviewer (MH) using a piloted form and checked by a second reviewer (KB) for correctness and completeness of the extracted data. The data extraction form included the categories generic bibliographic information, study characteristics, study objectives, hypothesis and research questions, population, and context characteristics, as well as findings and results of the critical appraisal (see Supplementary File: Appendix B). Data extraction for this systematic review considered the primary studies, and the modelling studies included.

Primary outcomes were incidence risk of SARS-CoV-2 infections among migrant and forcibly displaced populations, and outcomes of infections (e.g. measured by hospitalisation, admissions to intensive care units (ICU), or mortality). Where reported, outcomes were extracted by subgroups (e.g. by nationality, or contextual information such as accommodation type). Secondary outcomes were the effects of lockdown measures on the health status of refugees, asylum seekers, IDPs and migrants, especially referring to mental health outcomes.

### 2.4 Evidence Synthesis and Statistical analysis

Extracted data was tabulated and summarised by narrative synthesis and, where applicable, by statistical meta-analysis. Numbers of SARS-CoV-2 cases and respective population size, including data on sub-groups, were visualised in a forest plot along with corresponding 95% confidence intervals calculated by the ‘metaprop’ command in STATA SE 15 (with the “nooverall” option to supress pooling of studies across subgroups due to high heterogeneity).[14]

## 3. Results

The searches resulted in a total of 973 hits, and after duplicate removal, 715 studies remained for screening. Of these, 133 records were included for full-text screening and 13 met all the inclusion criteria. Using the snowballing method, two further studies meeting the inclusion criteria were detected. In total, 15 studies were included in this review (Figure 1).

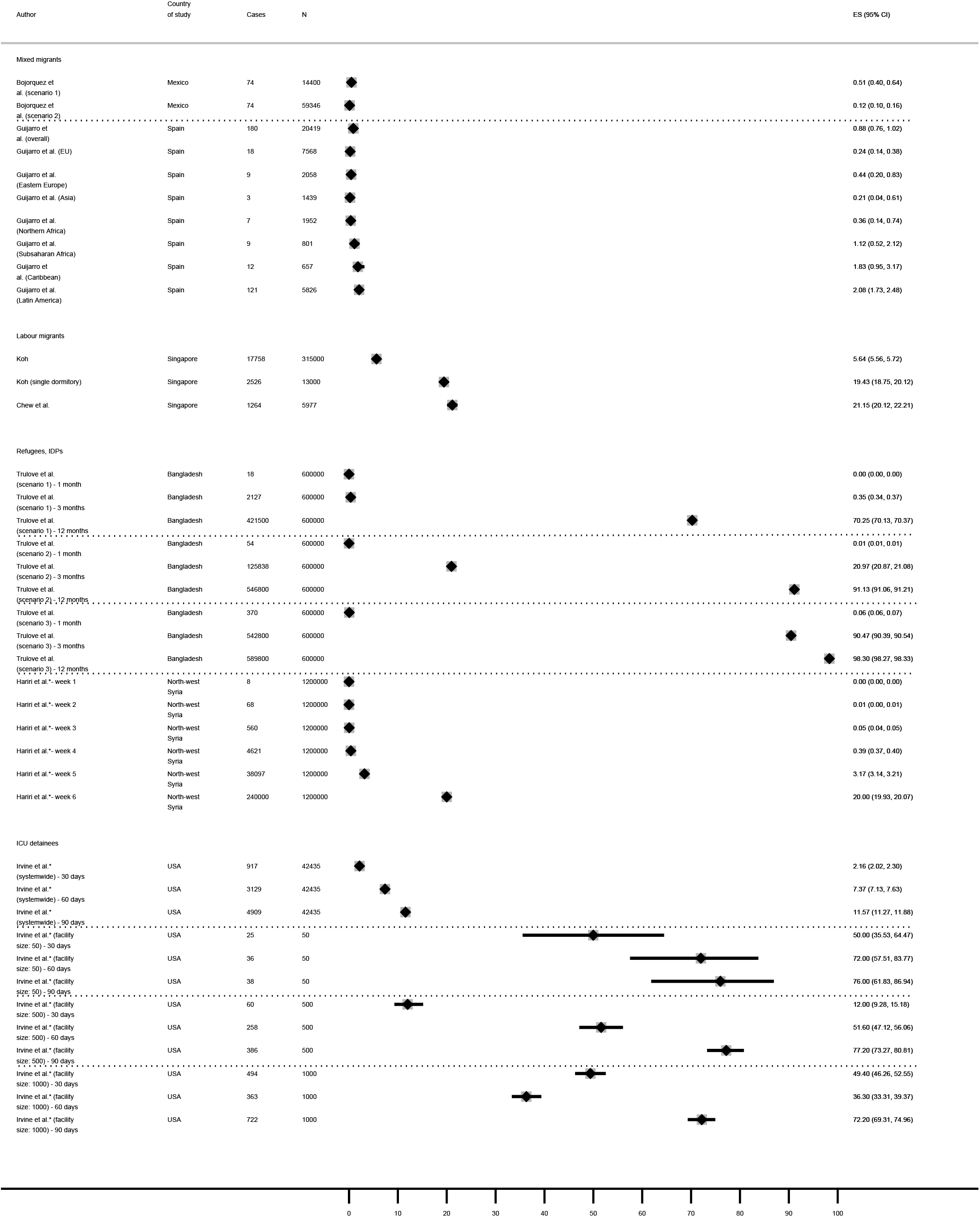

Quality appraisal using the EPHPP tool resulted in two studies at low risk of bias, four studies at moderate risk of bias, and six studies at high risk of bias. Appraisal of the modelling studies resulted in two studies at low risk of bias and one at high risk of bias. An overview of the quality appraisal is provided in the supplements [see detailed risk of bias assessment: Appendix C].

Almost half of the studies (n=7) reported data from high-income countries (HIC), five studies reported from upper or low middle-income countries (MIC), and three studies reported from low-income countries (LIC). The studies covered the following migrant and forcibly displaced population groups: refugees, asylum seekers or IDPs (n=5), migrant workers (n=4), international students (n=2), and migrants with no further specification (n=2). The reported outcomes were incidence risks among migrant population groups, modelled transmission scenarios, physical health outcomes such as hospitalisation, ICU admissions, or mortality, and mental or social impact of the COVID-19 pandemic on peoples’ health or well-being.

### 3.1 Incidence risk

The incidence risk among the observational studies varied from 0·12% to 2·08% in non-outbreak settings [16, 17], and from 5·64% to 21·15% in outbreak settings [18, 19], showing a high heterogeneity across the studies. Modelling studies also showed a high heterogeneity, which is due to different scenarios and timeframes. Detailed estimates on migrants and forcibly displaced populations are plotted in figure 2.

**Figure 2:**
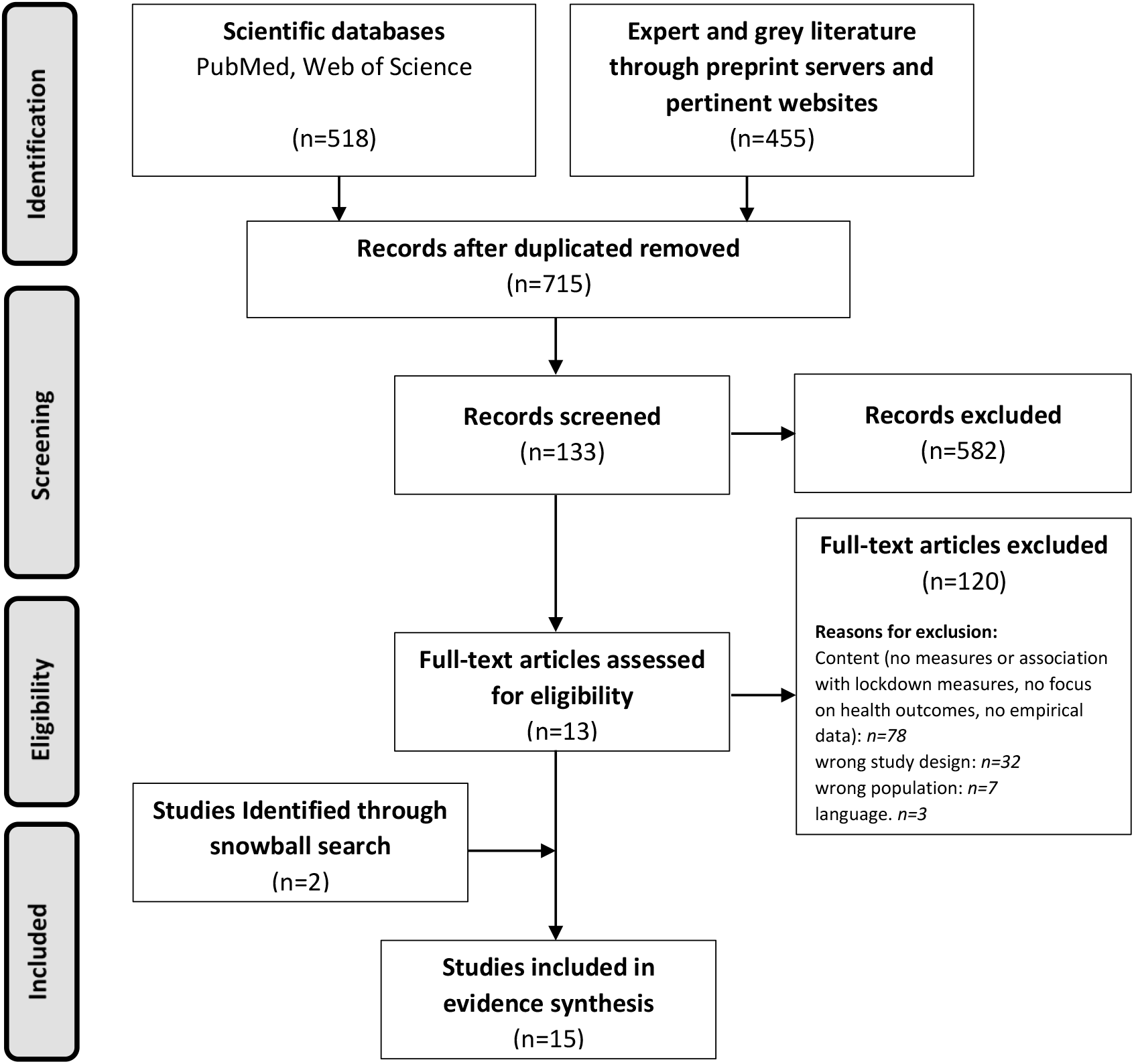
PRISMA Flow-Chart.

**Figure 2: Forest plot of Incidence risks**

**[see attached]**

Legend: Bojorquez et al. (szenario 1): Population of asylum seekers waiting to submit their asylum application in April 2020 [20]; Bojorquez et al. (szenario 2): Population refers to the number of cases in the United States immigration courts assigned to the MPP program from March 2019 to March 2020 [21]; Trulove et al. (scenario 1): Basic reproduction number R0= 1·5–2·0; Trulove et al. (scenario 2): Basic reproduction number R0= 2·0-3·0; Trulove et al. (scenario 3): Basic reproduction number R0= 3·3-5·0; Hariri et al*: Camp-population scenario with a very fast doubling rate of 2·3 days; Irvine et al.*: Only data from most optimistic scenario extracted (R0=2·5). Y-axis: percentage (%). ES: estimate of incidence proportion.

Studies with patient populations[17] report lower incidence rates compared to populations in congested settings [18, 19, 22-24]. However, the incidence risk varies among the different regions of origin of those infected (0·21-2·08%).[17] Compared to the incidence risk for the Spanish estimated at 0·7%, significantly higher rates were reported by a pre-print study for individuals from Sub-Saharan-Africa, Caribbean, and Latin America.[17] A high risk of bias study among migrants in transit in Mexico reports low incidence risks in two scenarios with 0·12% or 0·51% respectively.[16] The estimated cumulative incidence in two scenarios (with different sources for the population size of asylum seekers in the US) showed a higher number of suspected cases per 100 000 in the migrant group compared to non-migrants.[17] An ecological study investigating migrants in transit in Mexico found a higher Incidence Risk Ratio (IRR: 6·43, 95%CI, 4·41-9·39) for SARS-CoV-2 in state cluster populations with a higher proportion of migrants.[25] Contextual factors, such as dormitories, were associated with incidence risks in migrant workers in the range of 5·64% (95%CI, 5·56-5·72) to 19·43% (95%CI, 18·75-20·12)[18] and 21·15% (95%CI, 20·12-22·21)[19].

Another study assessed SARS-CoV-2 screening campaigns in different shelters and accommodation centres of homeless people and asylum seekers in Marseille, France. No SARS-CoV-2 cases were found among the asylum seekers tested on a voluntary basis, while among the 411 homeless people, facing similar living conditions, tested in different shelters 9% (n=37) were tested positive.[26]

### 3.2 Transmission Dynamics and Hospitalisation

Transmission scenarios were found to be slow in the beginning with a rapid increase of infections starting between the fourth week and the third month after start of simulated outbreaks.[22, 23] Irvine et al. found the speed of transmission in confined detention centres to be dependent on the facility size, starting with five infected individuals at baseline, where smaller facilities of 50 detainees reach the peak of infections earlier (day 19) than larger ones of 1000 detainees (day 69), assuming a moderate reproduction number of R(0)=3·5.[24] This may be emphasised by the incidence risks of the modelling studies [22-24] plotted in figure 2.

Four studies assessed hospitalisation among the respective populations. Irvine et al. modelled hospitalisations in three different scenarios with reproduction numbers between R(0)=2·5 and R(0)=7·0.[24] For the most pessimistic scenario (R(0)=7), models show that 15·1% of the Immigration and Customs Enforcement (ICE) detainees would need hospitalisation. For a facility housing 1000 detainees, this would mean a range of hospitalisation numbers between 10 and 117 at day 30 and between 114 and 157 at day 90 across all scenarios.[24] Truelove et al. estimated 4·8% (95%PI, 0·3%-15%) of 600.000 Rohingya refugees living in a camp to be severe cases and in need of hospitalisation across the transmission scenarios.[23] The authors calculated with reproduction rates between R(0)=1·5 and R(0)=5·0, also reaching up to 15% of hospitalisations in the worst case scenario.[23] The model also computed the point in time when needs exceed the hospital capacity at the Kutupalong-Balukhali Expansion Site in Bangladesh. In the low transmission scenario, this is the case after 136 days (95%PI, 96-196 days), and in the high transmission scenario after 55 days (95%PI, 42-77 days.).[23] Among the observational studies Bojorquez et al. found a tendency of lower odds to be hospitalised in migrants in transit and asylum seekers in Mexico (adjusted OR for confirmed cases: 0·18 (CI95%, 0·2-1·53) compared to the Mexican population.[16] In contrast, the odds of interstate migrants was 1·36 (95%CI, 1·19-1·54) times the odds of hospitalisation than the reference (Mexican) population.[25] About 6·49% of all hospitalised SARS-CoV-2 cases in Mexico from 28th February until 21st April 2020 were interstate migrants.[25] However, the IRR for hospitalisation in state cluster populations with a higher proportion of migrants showed an inverse association (IRR: 0·65, 95%CI 0·58-0·74).[25] A study among patient populations in Spain reports a 3-, 6-, and 7-fold higher risk among patients from Sub-Saharan-Africa, the Caribbean, and Latin America, respectively, of being PCR confirmed cases or being hospitalised compared to the Spanish patients.[17]

### 3.3 ICU admission

ICU admission was only reported by modelling studies. Irvine et al. found approximately 1·8% of the ICE detainee-population in need of ICU after 90 days of detention assuming a low transmission scenario R(0)=2·5, and approximately 2·3% within the same time within a high transmission scenario R(0)=7.[24] In comparison, the simulations of Trulove et al. result in lower estimates for ICU transmission in Rohingya refugees after three months’ time, while using lower reproduction rates.[23] Cumulative numbers of ICU admissions are ranging from 6 (0-30) to 4 440 (98-10 100) across the scenarios, which accounts for 0·0%-0·7% of the Rohingya refugee population.[23] Hariri and colleagues calculate with a doubling rate of 2·3 leading to 12 000 (1%) critical cases in need of ICU admission among IDPs in camps and tented settlements along the Turkey-Syria border.[22] Moreover, their scenario estimates a 50% overload of ICU capacity within the fourth week.[22] Only one study used empirical instead of modelling data to investigate the need of intubation within a patient population and reports that 12·5% of migrants need intubation. However, due to a high number of missing values, these results are to be interpreted with caution.[16] Risk conditions in this specific study population seemed to be less among migrants except for the case of pregnant women.[16]

### 3.4 Mortality

Five studies address mortality as an outcome variable. Proportions of migrants with a fatal outcome in the studied populations vary from 0·48% to 6·0% across the studies. The modelling study of Truelove et al. shows the lowest fatality rate with 0·48% (95%PI, 0·03%-1·5%) assuming that 10% of severe cases result in death.[23] The model of Hariri and colleagues predicts 1·6% (n= 18 751) deaths within the first six weeks upon infection.[22] The authors mention that numbers might be higher due to a lack of health facility capacity in northwest Syria for the treatment of severe and critical cases.[22] Non-modelling studies report a mortality rate among interstate migrants of 4·75% (95%CI, 4·06-5·43) with an odds ratio for fatal outcomes of 2·01 (95%CI, 1·46-2·76) compared to average state-cluster characteristics.[25] The IRR for lethality in state-clusters with higher proportions of migrants (IRR: 1·02, 95%CI, 0·98-1·02) showed a mild tendency to be higher in these populations.[25] Guijarro et al. investigate SARS-CoV-2 cases at a hospital in Alcorcón, Spain, finding 33% of severe cases in migrants (defined by country of origin) compared to 63% in the Spanish population.[17] Severe cases include death, critical care admission, and hospital stay longer than 7 days. Unadjusted mortality among severe cases was at 6% in the migrant and 25% among the Spanish population.[17] Motta et al. also found a lower mortality rate in migrants (2·3%, n=1, N=43) compared to non-migrants (26·9%, n=7, N=26) in the investigated study population of tuberculosis patients infected with SARS-CoV-2.[27]

### 3.5 Mental health outcomes and well-being

Mental and social wellbeing among different migrant groups was assessed in five studies. Kumar and colleagues found 73·5% out of 98 migrant workers in India to be screened positive for depression, anxiety, and/or perceived stress due to pandemic and lockdown measures measured by PHQ2, GAD2 and PSS-4 questionnaires.[28] In line with that, Qiu et al. assessed psychological levels of distress among Chinese people during the COVID-19 pandemic finding the highest level of distress among migrant workers compared to other, non-migratory occupation groups.[29]

Impact on social and socio-psychological wellbeing was assessed among international students in central China’s Hubei province, showing that almost a quarter of the study population talked about the virus in regular conversations and almost half of the study population was unhappy due to the current lockdown condition.[30] Moreover, migrant workers in India reported a deterioration of social-wellbeing with 63·3% reporting increased loneliness and about half of the participants indicating a significant increase in negative feelings such as tension, frustration, irritability, and fear of death.[28] Reduction of sleep and social connectedness affected about one third of the participating migrant workers in the same study.[28] Another topic investigated by Lopez-Pena and colleagues were health behaviours of Rohingya refugees during the COVID-19 pandemic.[31] The authors focused on health providers chosen by persons that showed symptoms of SARS-CoV-2 and at trusted information sources such as friends, newspapers or non-governmental organisations (NGO). A total of 42·3 % (95%CI 32·4-52·3) of symptomatic household members in refugee camps chose pharmacies as their first-choice health provider followed by health information providers in camps (35·8% (95%CI 26·0-45·6)).[31] Trusted information sources on COVID-19 prevention and advice among Rohingya refugees were friends, neighbours, and acquaintances (58·8% (95%CI 50·7-66·9)) followed by NGOs (53·5% (95%CI 45·6-61·3)) and informational campaigns on the streets (41·6% (95%CI 33·6-49·7)).[31]

Rzymski and colleagues assessed prejudices against Chinese students studying at a university in Poland to assess social wellbeing.[32] According to the survey, 61·2% of participants said that they experienced prejudices in public transport, shopping or in restaurants and in health services due to the COVID-19 pandemic.[32] Racist behaviours were shown in preconceptions and rejective behaviours of others against Chinese international students resulting in negative impacts on their wellbeing.[32]

## 4. Discussion

This systematic review has demonstrated a high heterogeneity of health consequences of the COVID-19 pandemic in migrant and forcibly displaced populations, who are in turn a heterogenous group exposed to a wide range of living conditions. Compared to various non-migrant reference groups, the incidence risks reported among migrant and forcibly displaced populations tend to be consistently higher, while hospitalisation rates and ICU admissions seem to be lower among migrants. However, most findings for the latter are derived from modelling studies and may thus only approximate reality as, e.g. hospital bed and ICU capacity were not always considered. As expected based on knowledge from studies in general populations, the different transmission scenarios in modelling studies show an exponential increase in SARS-CoV-2 cases after the start of transmission within a migrant population. There was no substantial gain in evidence regarding mortality rates as results were mixed. Crude mortality rates derived from observational studies were higher among patient populations compared to population-based studies, but overall were lower among migrants compared to non-migrant populations. In contrast, a population-based study at moderate risk of bias found a higher lethality in populations with a higher proportion of migrants when adjusted for age and gender. All migrant and forcibly displaces populations investigated by the included studies, were exposed to precarious living and working conditions and hence were at a higher risk of becoming infected or severe cases. In addition, the evidence consistently suggests that mental health is negatively impacted by the pandemic situation across all migrant groups considered by this review. Apart from the reduction in social connectedness, migrant workers no longer have job security and are therefore strongly affected by lockdown measures. However, as there is a lack of comparative studies assessing whether migrants and forcibly displaced populations suffer to a different extent from the pandemic measures compared to native reference populations, generalised conclusions cannot yet be drawn in this respect.

Overall, we found a heterogeneous and fragmented research landscape studying the health of migrants and forcibly displaced populations during the COVID-19 pandemic, few high-quality studies, and a scarcity of comparative designs.

There is a need for more analytical studies using robust comparative study design to monitor inequalities regarding exposure to SARS-CoV-2 or related pandemic policies. This requires a strengthening of health information systems, that however, due to weak capacities, are falling short of instantly generating and providing health information data on migrants, asylum seekers, refugees and IDPs.[33] Moreover, no qualitative studies were available at the time our search was conducted. Many researchers and organisations already called for the integration of vulnerable population groups such as refugees and migrants into national policy plans [34-36] and qualitative studies are instrumental to provide insights into realities of migrant and forcibly displaced population groups during the pandemic. At structural level, and as long as medical remedies are absent, improving living conditions of these groups by ensuring preconditions that allow for self-isolation, physical distancing, and hygiene[37] appear to be the best preventive policy measures to protect migrants and forcibly displaced populations from being particularly exposed to SARS-CoV-2.

### 4.1 Strengths and limitations

To our knowledge, and beyond a bibliometric analysis of SARS-CoV-2 research and migration[38], this is the first review to investigate empirical data available on migrant populations at this stage of the COVID-19 pandemic.

The conception of this review as a rapid systematic review made it possible to conduct search, screening, quality appraisal, data extraction and synthesis in a timely manner. At the same time, we had to compromise by restricting the study languages to English and German, which poses a possible limitation to identify all empirical data available on this topic so far. The heterogeneity of studies did not allow for running a meta-analysis with pooled estimates to gain further knowledge about the incidence risk in migrant populations.

Furthermore, the body of evidence included is limited by a scarcity of high-quality studies and prone to a wide range of bias (hospital bias, diagnostic bias, selection bias, and misclassification bias) or residual confounding. Mortality studies, for example, did not always adjust for age and comorbidity when comparing migrants and non-migrants (see detailed risk of bias assessment: Appendix C). The inclusion of pre-prints, comments, or letters to the editor reporting empirical data was also a challenge for quality appraisal. Nevertheless, this was necessary in order to find as much empirical data as possible, at the early stage of the pandemic when our search was conducted. Given the dynamic number of SARS-CoV-2 related publications, updates of the rapid review will be required in regular intervals to synthesise and consider emerging evidence.

## 4.2 Conclusion

The summarised evidence in this first systematic review on SARS-CoV-2 among migrant and forcibly displaced populations shows high incidence risks among migrants, refugees, asylum seekers and IDPs, yet low hospital admission rates, and mixed mortality-related results. Due to the tenuous and heterogenous data situation on which the review is based on, results need to be interpreted with caution. In view of the general scarcity of health data on migrant and forcibly displaced populations, the pandemic might rather be a barrier than a facilitator to improve the body of evidence. More robust and comparative study designs are urgently needed to assess differences and inequalities in risk of infection, consequences of disease, contextual risk factors, and impact of pandemic policies among migrants and forcibly displaced populations. This might include strengthening of health information systems and integration of these groups in notification and reporting systems at all levels of the healthcare system.

## Data Availability

All Data referred to in the Manuscript is already published either as peer-reviewed article, comment, letter to the editor or as pre-print.

## Acknowledgments

None.

## Author Contributions

Search: Pubmed and WoS: SR; preprint servers: MH; Websites: MT and KK

Title and abstract screening: HG, MH

Full-text screening: HG, MH, JS, CSH, KB

Quality appraisal: MH, KB

Dataextraction: MH, KB

Data-synthesis: MH, KB

Writing of first and final draft: MH, KB

Revision for important intellectual content: JS, MT, SR, HG, KK, C-SH

## Ethics statement

The study is based on published literature, no ethical clearance was required.

## References

1. Bozorgmehr K, Saint V, Kaasch A et al. (2020) COVID and the convergence of three crises in Europe. The Lancet Public Health 5(5):e247–e248

2. Schenker MB (2010) A global perspective of migration and occupational health. American journal of industrial medicine 53(4):329–337

3. Organization WH (2020) Report of the WHO-China joint mission on coronavirus disease 2019 (COVID-19). Geneva

4. Hargreaves S, Rustage K, Nellums LB et al. (2019) Occupational health outcomes among international migrant workers: a systematic review and meta-analysis. The Lancet Global Health 7(7):e872–e882

5. Lancet T (2020) India under COVID-19 lockdown. Lancet (London, England) 395(10233):1315

6. Sze S, Pan D, Nevill CR et al. (2020) Ethnicity and clinical outcomes in COVID-19: A systematic review and meta-analysis. EClinicalMedicine:100630-100630

7. Tai DBG, Shah A, Doubeni CA et al. (2020) The disproportionate impact of COVID-19 on racial and ethnic minorities in the United States. Clinical Infectious Diseases

8. Garritty C, Gartlehner G, Kamel C et al. (2020) Cochrane rapid reviews. Interim guidance from the Cochrane rapid reviews methods Group. March

9. Hintermeier M, Kajikhina K, Rohleder S et al. (2020) Refugees and migrants and COVID-19: a rapid systematic review. PROSPERO (CRD42020195633)

10. Bennett C, Manuel DG (2012) Reporting guidelines for modelling studies. BMC medical research methodology 12(1):168

11. Saltelli A, Bammer G, Bruno I et al. (2020) Five ways to ensure that models serve society: a manifesto. Nature Publishing Group

12. Egger M, Johnson L, Althaus C et al. (2017) Developing WHO guidelines: time to formally include evidence from mathematical modelling studies. F1000Research 6

13. Dahabreh IJ, Trikalinos TA, Balk EM et al. (2016) Guidance for the conduct and reporting of modeling and simulation studies in the context of health technology assessment Methods Guide for Effectiveness and Comparative Effectiveness Reviews. Agency for Healthcare Research and Quality (US)

14. Nyaga, Nyaga V, Arbyn M et al. (2014) Metaprop: a Stata command to perform meta-analysis of binomial data. Archives of Public Health 72

15. (2020) Competence Network Public Health Covid-19 https://www.public-health-covid19.de/en/ (last accessed: 30.11.2020)

16. Bojorquez I, Infante C, Vieitez I et al. (2020) MIGRANTS IN TRANSIT AND ASYLUM SEEKERS IN MEXICO: AN EPIDEMIOLOGICAL ANALYSIS OF THE COVID-19 PANDEMIC. medRxiv:2020.2005.2008.20095604

17. Guijarro C, Perez-Fernandez E, Gonzalez-Pineiro B et al. (2020) Risk for COVID-19 among Migrants from different areas of the world in Spain: A population-based cohort study in a country with universal health coverage. medRxiv:2020.2005.2025.20112185

18. Koh D (2020) Migrant workers and COVID-19. Occupational and environmental medicine

19. Chew MH, Koh FH, Wu JT et al. (2020) Clinical assessment of COVID-19 outbreak among migrant workers residing in a large dormitory in Singapore. The Journal of hospital infection

20. Leutert S, Arvey S, Ezzell E et al. (2020) Metering & COVID-19

21. TRAC (2019) Details on MPP (Remain in Mexico) Deportation Proceedings

22. Hariri M, Rihawi H, Safadi S et al. (2020) THE COVID-19 FORECAST IN NORTHWEST SYRIA The Imperative of Global Action to Avoid Catastrophe. medRxiv:2020.2005.2007.20085365

23. Truelove S, Abrahim O, Altare C et al. (2020) The potential impact of COVID-19 in refugee camps in Bangladesh and beyond: A modeling study. PLoS medicine 17(6):e1003144

24. Irvine M, Coombs D, Skarha J et al. (2020) Modeling COVID-19 and Its Impacts on U.S. Immigration and Customs Enforcement (ICE) Detention Facilities, 2020. Journal of urban health : bulletin of the New York Academy of Medicine:1–9

25. Mendez-Dominguez N, Alvarez-Baeza A, Carrillo G (2020) Demographic and Health Indicators in Correlation to Interstate Variability of Incidence, Confirmation, Hospitalization, and Lethality in Mexico: Preliminary Analysis from Imported and Community Acquired Cases during COVID-19 Outbreak. International journal of environmental research and public health 17(12)

26. Ly TDA, van Hoang T, Goumballa N et al. (2020) Screening of SARS-CoV-2 among homeless people, asylum seekers and other people living in precarious conditions in Marseille, France, March April 2020. medRxiv:2020.2005.2005.20091934

27. Motta I, Centis R, D’Ambrosio L et al. (2020) Tuberculosis, COVID-19 and migrants: preliminary analysis of deaths occurring in 69 patients from two cohorts. Pulmonology

28. Kumar K, Mehra A, Sahoo S et al. (2020) The psychological impact of COVID-19 pandemic and lockdown on the migrant workers: A cross-sectional survey. Asian J Psychiatr 53:102252

29. Qiu J, Shen B, Zhao M et al. (2020) A nationwide survey of psychological distress among Chinese people in the COVID-19 epidemic: implications and policy recommendations. General psychiatry 33(2)

30. Fakhar-e-Alam Kulyar M, Bhutta ZA, Shabbir S et al. (2020) Psychosocial impact of COVID-19 outbreak on international students living in Hubei province, China. Travel Medicine and Infectious Disease:101712

31. Lopez-Pena P, Davis CA, Mobarak AM et al. (2020) Prevalence of COVID-19 symptoms, risk factors, and health behaviors in host and refugee communities in Cox’s Bazar: A representative panel study. Bull World Health Organ

32. Rzymski P, Nowicki M (2020) COVID-19-related prejudice toward Asian medical students. Journal of Infection and Public Health 13(6):873–876

33. Organization WH (2019) Health Evidence Network synthesis report 66: what is the evidence on availability and integration of refugee and migrant health data in health information systems in the WHO European region?

34. Bhopal RS (2020) COVID-19: Immense necessity and challenges in meeting the needs of minorities, especially asylum seekers and undocumented migrants. Public Health 182:161

35. Lancet T (2020) COVID-19 will not leave behind refugees and migrants. Lancet (London, England) 395(10230):1090

36. Tallarek M, Bozorgmehr K, Spallek J (2020) Toward inclusionary and diversity-sensitive public health: the consequences of exclusionary othering in public health using the example of COVID-19 management in German reception centers and asylum camps. BMJ Global Health:In Press

37. Orcutt M, Patel P, Burns R et al. (2020) Global call to action for inclusion of migrants and refugees in the COVID-19 response. Lancet (London, England) 395(10235):1482–1483

38. Pernitez-Agan S, Bautista MA, Lopez J et al. (2020) Bibliometric Analysis of COVID-19 in the Context of Migration Health: A Study Protocol. medRxiv:2020.2007.2009.20149401

