## APPENDIX_A for "SARS-CoV-2 among migrants and forcibly displaced populations: a rapid systematic review": APPENDIX_A_search_queries.pdf

### Search Queries in PubMed, WoS, Pre-print Servers and Website search

| Search Syntax development (MEDLINE via PubMed) on 26 June 2020 | | |
|---|---|---|
| **ID** | **Query** | **Results** |
| **#11** | **#4 AND #9 AND #10** | **346** |
| #10 | 2019/12/01:2020/06/26[edat] | 759,959 |
| #9 | #5 OR #6 OR #7 OR #8 | 845,962 |
| #8 | refugee[tw] OR “asylum seeker”[tw] OR “asyl*”[tw] OR (displaced[tw] AND (person*[tw] OR people[tw])) OR “migra*”[tw] OR “forced migra*”[tw] OR migration[tw] OR immigra*[tw] OR “human migration”[tw] OR stateless[tw] OR “state-less”[tw] OR “irregular migra*”[tw] OR “regular migra*”[tw] OR “undocumented migra*”[tw] OR “internally displaced”[tw] OR “detainees”[tw] OR “residence status”[tw] OR “foreign-born”[tw] OR “ displaced person”[tw] OR “noncitizen”[tw] OR “outsider”[tw] OR “newcomer”[tw] OR “newly arrived”[tw] OR “new arrival”[tw] OR “recent entrant”[tw] OR “non national”[tw] OR “non-national”[tw] OR “transient”[tw] OR “minorities”[tw] OR “ethnic”[tw] | 842,865 |
| #7 | “Refugees”[MeSH] | 10,151 |
| #6 | “Human Migration”[MeSH] | 26,250 |
| #5 | “Transients and Migrants”[MeSH] | 11,563 |
| #4 | #1 OR #2 OR #3 | 41,990 |
| #3 | ("corona virus"[tw] OR "corona viruses"[tw] OR coronavir*[tw] OR coronovirus*[tw] OR betacoronavirus*[tw]) AND (novel[tw] OR 2019[tw] OR Wuhan[tw] OR Huanan[tw] OR Hubei[tw]) OR "new coronavirus"[tw] OR "COVID-19"[tw] OR COVID19[tw] OR "SARS coronavirus 2"[tw] OR "severe acute respiratory syndrome coronavirus 2"[tw] OR nCoV[tw] OR 2019nCoV[tw] OR nCoV2019[tw] OR "SARS-CoV-2"[tw] OR "SARS-CoV2"[tw] OR SARSCoV19[tw] OR SARS-CoV19[tw] OR SARS-CoV-19[tw] OR HCoV-19[tw] OR WN-CoV[tw] | 28,644 |
| #2 | "Coronavirus Infections"[Mesh] | 17,903 |
| #1 | "Coronavirus"[Mesh] | 18,272 |

| Search Syntax development (Web of Science Core Collection) on 26 June 2020 | | |
|---|---|---|
| **ID** | **Query** | **Results** |
| #4 | #1 AND #2 Timespan=2019-2020 | 172 |
| #3 | #1 AND #2 | 405 |
| #2 | TS = (refugee OR "asylum seeker" OR "asyl*" OR (displaced AND (person* OR people) ) OR "migra*" OR "forced migra*" OR migration OR immigra* OR "human migration" OR stateless OR "state-less" OR "irregular migra*" OR "regular migra*" OR "undocumented migra*" OR "internally displaced" OR "detainees" OR "residence status" OR "foreign-born" OR " displaced person" OR "noncitizen" OR "outsider" OR "newcomer" OR "newly arrived" OR "new arrival" OR "recent entrant" OR "non national" OR "non-national" OR "transient" OR "minorities" OR "ethnic") | 1,339,608 |
| #1 | TS = (("corona virus" OR "corona viruses" OR coronavir* OR coronovirus* OR betacoronavirus*) AND (novel OR 2019 OR Wuhan OR Huanan OR Hubei) OR "new coronavirus" OR "COVID-19" OR COVID19 OR "SARS coronavirus 2" OR "severe acute respiratory syndrome coronavirus 2" OR nCoV OR 2019nCoV OR nCoV2019 OR "SARS-CoV-2" OR "SARS-CoV2" OR SARSCoV19 OR SARS-CoV19 OR SARS-CoV-19 OR HCoV-19 OR WN-CoV OR “corona vir*” OR “coronavir*” OR “betacoronavir*” OR “severe acute respiratory syndrome coronavirus”) | 26,076 |

| Seaches in Pre-print servers and websites | | | |
|---|---|---|---|
| **Source** | **Search expression** | **Hits** | **Date** |
| medRxiv via https://www.medrxiv.org/ | ""refugee" OR "migrants" OR "asylum seeker" OR "IDP"" and posted between "01 Dec, 2019 and 26 Jun, 2020" | 187 | 26 June 2020 |
| medRixiv via https://www.medrxiv.org/ | for term ""refugee" OR "migrants" OR "asylum seeker" OR "IDP" AND "COVID-19"" and posted between "01 Dec, 2019 and 26 Jun, 2020" | 6 | 26 June 2020 |
| bioRxiv via https://www.medrxiv.org/ | """refugee" OR "migrants" OR "asylum seeker" OR "IDP"" AND "COVID-19"" and posted between "01 Dec, 2019 and 26 Jun, 2020" | 4 | 26 June 2020 |
| WHO COVID research https://search.bvsalud.org/global-literature-on-novel-coronavirus-2019-ncov/advanced/?lang=en | tw:( (tw:(migra*)) OR (tw:(refugee*)) OR (tw:(idp))) AND la:("en" OR "de") AND (year_cluster:[2019 TO 2020]) | 174 | 29 June 2020 |
| EUPHA https://eupha.org/recent-news; https://eupha.org/covid---19-updates | Website search function (strg+f, search terms "refugee" OR "migrant" OR "asylum" OR "displaced" OR "forced migration" OR "migration" OR "migra" OR "regular" OR "irregular" OR "undocumented" OR "foreign" OR "international" OR "ethnic" OR "minorit*" OR "stateless" OR "residen*" OR "non-citizen") and key words for COVID-19/SARS-CoV2 exposure (see above), manual check for relevant documents | 13 | 25 June 2020 |
| IOM https://migrationhealthresearch.iom.int/; https://gmdac.iom.int/: | Website search function (strg+f, search terms "refugee" OR "migrant" OR "asylum" OR "displaced" OR "forced migration" OR "migration" OR "migra*" OR "regular" OR "irregular" OR "undocumented" OR "foreign" OR "international" OR "ethnic" OR "minorit*" OR "stateless" OR "residen*" OR "non-citizen") and key words for exposure (see above), manual check for relevant documents | 64 | 25 June 2020 |
| NIPH Live map COVID-19 evidence: https://www.nornesk.no/forskningskart/NIPH_mainMap.html | Search filter: "ethnicity" (no results in other categories refugee camps, other, other groups) | 7 | 25 June and 29 June 2020 |
