## APPENDIX_B for "SARS-CoV-2 among migrants and forcibly displaced populations: a rapid systematic review": APPENDIX_B_dataextraction_file.pdf

**APPENDIX B**
**Data Extraction file**

| Generic Bibliographic Information | | | Study Characteristics |
|---|---|---|---|
| **Author(s)** | **Date of Publication** | **Type of Publication** | **Type of Study** |
| M. Irvine, D. Coombs, J. Skarha, B. Del Pozo, J. Rich, F. Taxman and T. C. Green | 2020 | peer-reviewed | quantitative, modeling study |
| S. Truelove, O. Abrahim, C. Altare, S. A. Lauer, K. H. Grantz, A. S. Azman and P. Spiegel | 2020 | peer-reviewed | quantitative, modeling study |

| | | | |
|---|---|---|---|
| M. Hariri, H. Rihawi, S. Safadi, M. A. McGlasson and W. Obaid | 2020 | pre print | quantitative, modeling study |
| I. Bojorquez, C. Infante, I. Vieitez, S. Larrea and C. Santoro | 13 May 2020 | pre-print | quantitative, secondary analysis |

| | | | |
|---|---|---|---|
| K. Kumar, A. Mehra, S. Sahoo, R. Nehra and S. Grover | 2020 | letter to the editor | cross-sectional survey |
| D. Koh | 2020 | short report | - |

| | | | |
|---|---|---|---|
| I. Motta, R. Centis, L. D'Ambrosio, J.-M. García-García, D. Goletti, G. Gualano, F. Lipani, F. Palmieri, A. Sánchez-Montalvá, E. Pontali, G. Sotgiu, A. Spanevello, C. Stochino, E. Tabernero, M. Tadolini, M. van den Boom, S. Villa, D. Visca and G. B. Migliori | 14 May 2020 | peer-reviewed | quantitative |
| Mendez-Dominguez, N; Alvarez-Baeza, A; Carrillo, G | 15 June 2020 | peer-reviewed | quantitative: epidemiologic, cross-sectional study |

| | | | |
|---|---|---|---|
| T. D. A. Ly, V. T. Hoang, N. Goumballa, M. Louni, N. Canard, T. L. Dao, H. Medkour, A. Borg, K. Bardy, V. Esteves-Vieira, V. Filosa, B. Davoust, O. Mediannikov, P.-E. Fournier, D. Raoult and P. Gautret | 14 May 2020 | pre-print | quantitative, cross sectional |
| Guijarro, C; Perez-Fernandez, E; Gonzalez-Pineiro, B; Melendez, V; Goyanes, MJ; Renilla, ME; Casas, ML; Sastre, I; Velasco, M | 27 May 2020 | pre-print | quantitative, |

| | | | |
|---|---|---|---|
| M. H. Chew, F. H. Koh, J. T. Wu, S. Ngaserin, A. Ng, B. C. Ong and V. J. Lee | 31 May, 2020 | letter to the editor | quantitative |
| P. Rzymski and M. Nowicki | 26. Apr 20 | peer-reviewed | quantitative, |
| Fakhar-e-Alam Kulyar, M; Bhutta, Zeeshan A; Shabbir, S; Akhtar, M | 25.04.2020 | letter to the editor | quantitative, cross sectional |

| | | | |
|---|---|---|---|
| Lopez-Pena P, Austin Davis C, Mushfiq Mobarak A & Raihan S | 11.05.2020 | pre-print | quantitative |
| Qiu, J., Shen, B., Zhao, M., Wang, Z., Xie, B., & Xu, Y | 29.02.2020 | Editorial | quantitative |

| Period of Study | Context of Study/Setting |
| --- | --- |
| october 1, 2019 - march 1, 2020 | U.S. Immigration and Customs Enforcement (ICE) facilities, where detained immigrants are not housed individually or in cells as is true in many prison settings (detainees have more contacts with one another and with staff, thereby contributing to faster spread of infection than in correctional facilities) |
| Not explicitly stated. | The Kutupalong-Balukhali Expansion Site, Bangadesh |

| | |
|---|---|
| Almost 1 million civilians were displaced by the escalation of hostilities between **December 2019 and March 2020** | camps and tented settlements along the Turkey-Syria border |
| January 1, 2020 - May 3, 2020 | migrants in transit and asylum seekers in Mexico |

| | |
|---|---|
| The data collection was done during the 5th week of lockdown with adherence to social distancing norms and other infection control measures. | Chandigarh, a Union Territory, in North India |
| Information was gathered from daily reports provided by the Ministry of Health, Singapore Statues online and a Ministerial statement given at a Parliament sitting on 4 May 2020 | Dormitory housing of migrant workers in Singapore |

| | |
|---|---|
| The first COVID-19 diagnosis was made on March 12th, 2020; data were updated as of May 5th, 2020. | 26 centres in Belgium, Brazil, France, Italy, Russia, Singapore, Spain, and Switzerland |
| from the beginning of the outbreak (Mexico: 28th of Februrary) to 21 April 2020 | Sentinel surveillance for COVID-19 cases in Mexico |

| | |
|---|---|
| 26 March - 17 April 2020 | different shelters and accomodation centres in Marseille, France |
| February 1 until April 25 2020 | Hospital Universitario Fundación Alcorcón, Spain |

| | |
|---|---|
| April 11th to April 19th, 2020 | dormitories in Singapore |
| February 2020 before the first case of COVID-19 was confirmed in Poland on March 4, 2020 | Poznan University of Medical Sciences in Poland |
| | international students living in Hubei province |

| | |
|---|---|
| 11. -17.April 2020 | phone based survey in households across Cox's Bazar in Bangladeshi and Rohingya Refugees living in camps in Cox's Bazar |
| start: 31.01.2020- 10.02.2020 | psychological distress in the general population of China during the tumultuous time of the COVID-19 epidemic |

| Research Objectives |
| --- |
| As COVID-19 continues to spread in communities, and inevitably into the nation’s detention centers, it is critical that we understand the US healthcare system’s capacity to care for a large influx of patients who require critical care. In the present study, we estimate the number of COVID-19 cases likely to occur within ICE detention facilities, using varying levels of virus Irvine et al. transmissibility, and examine the capacity of local hospitals to care for additional patient ICE detainees over 30-, 60-, and 90-day time horizons |
| The primary aims of this analysis are to (1) develop a baseline expectation of the possible infection burden, speed, and hospitalization capacity needed to respond to a COVID-19 epidemic; (2) use these findings to provide some recommendations to support ongoing preparedness planning by the Bangladesh government, United Nations agencies, and other actors for a COVID-19 outbreak; and (3) apply lessons from this case study to refugees and other forcibly displaced persons globally. |

| |
|---|
| The objectives are to: Generate estimates of projected case load, by severity, deaths and health system needs over the first 8 weeks of an epidemic according to various scenarios affecting the whole population or the highly-vulnerable population of camp dwellers; identify critical timepoints at which the health system capacity to manage COVID-19 cases is exceeded and the health system is at risk of collapse; and use these findings to draw recommendations to support ongoing preparedness planning |
| To describe the epidemiology of suspect cases of COVID-19 in migrants in transit and asylum seekers in Mexico, and to compare their characteristics with those of non-migrants |

| |
|---|
| Accordingly, this study aims to evaluate the mental health issues among the migrant workers living in shelter houses, provided by the administration because of COVID-19 to assess the immediate and long term psychological impact of isolation. |
| Daily numbers of COVID-19 in Singapore from March to May 2020, the cause of a surge in cases in April and the national response were examined, and regulations on migrant worker accommodation studied. |

| |
|---|
| The aim of this study is to describe for the first time a group of patients who died with TB (active disease or sequelae) and COVID-19 in the cohorts A and B. |

This study’s objective is to analyze the incidence, lethality, hospitalization, and confirmation of COVID-19 cases in Mexico

In this study, we present the results of SARS-CoV-2 screening campaigns conducted among sheltered homeless individuals, in comparison with asylum-seekers, other persons living in precarious conditions, and employees working in the accommodation centres. We also investigated the role of potential risk factors for virus carriage among the homeless population.

We sought to describe the incidence of COVID-19 among migrants from different areas of the world as compared to Spaniards, both of them living in Alcorcón, a city in the suburbs of Madrid (Spain) with a substantial proportion of foreign residents. The Spanish Health System provides universal free access to medical care for all inhabitants through the Madrid’s Health Service (SERMAS).

| |
|---|
| Dormitory A is a large privately owned dormitory that can accommodate 13,000 inhabitants, in rooms of 12 residents across 13 blocks. With the high number of COVID-19 cases arising from dormitory A, it was gazetted by law as an isolation area on April 5th [2]. We describe results of the outbreak investigation and clinical evaluation in dormitory A. |
| To identify whether individuals of Asian origin may experience any prejudice related to SARS-CoV-2, we have conducted an anonymous online survey of Asian medicine students at Poznan University of Medical Sciences in Poland. |

Our aim of study was to explore that panic and influencing factors on such students

| |
|---|
| We study the prevalence of common symptoms of COVID-19 and its correlates among Rohingya refugee camps and Bangladeshi host communities in Cox's Bazar. The district is currently home to almost 860,000 stateless Rohingya refugees, the vast majority of whom reside in densely populated camps and depend on emergency aid to cover daily needs. We document how the prevalence of COVID-19 symptoms is associated with transmission vectors, compliance with various social distancing recommendations, and current and mid-2019 living conditions. We also describe trusted information sources of refugees and hosts, and the preferred healthcare providers of each group. |
| The main purpose of this study is to measure the prevalence and severity of this psychological distress, gauge the current mental health burden on society, and therefore provide a concrete basis for tailoring and implementing relevant mental health intervention policies to cope with this challenge efficiently and effectively |

| Research Hypothesis / assumptions for modeling studies |
|---|
| - To prevent stochastic burnout, we performed simulations using an initial condition of five infectious detainees.<br>- We used the minimum rate for each age group in our calculations.<br>**- Initial conditions**: On day 0 of the epidemic, five individuals are assumed to be infected (in state I), with the rest being susceptible for a range of fixed population sizes chosen to be reflective of facility sizes: 50, 100, 500, and 1000.<br>- Drawing from the published estimates, we studied three scenarios ranging from a most optimistic to most pessimistic estimate of the R0 as 2.5, 3.5, and 7 for low, medium, and high R0 scenarios, respectively. The incubation and infectious periods were estimated to be 6.4 days [16] and 7 days [17], respectively<br>- The average number of detainees between October 1, 2019 and March 2, 2020 was used to approximate the current number of people detained at each detention facility and summed to approximate the total number of people currently detained by ICE<br>- We determined ICU capacity using the Definitive Healthcare USA Hospital Bed data<br>- Radii of 10 and 50 miles were selected as healthcare service catchment areas within which detention facilities may reasonably be expected to transport for patient care |
| - To capture the potential variability of transmission possible in this setting, we simulated epidemics under three potential scenarios with different values of the basic reproductive number, R0; The R0 in each of these scenarios falls within the 95% confidence interval (CI) of the current range of estimates for COVID-19<br>- We assumed an Erlang-distributed serial interval (time between the onset of symptoms in infector–infectee pairs) with a mean of 6 days (standard deviation [SD] = 4.2) [21].<br>- We assumed a population of 600,000 individuals and that the population was essentially closed (i.e., no movement, births, or deaths other than from COVID-19). Population characteristics and parameters used are detailed in Table 1, and further details about the model are in S1 Text.<br>- We assume that, in this setting, hospitalization would be limited to those with severe disease (defined as tachypnea [≥30 breaths/minute] or oxygen saturation ≤93% at rest, or PaO2/FIO2 <300, and/or lung infiltrates >50% of the lung field within 24–48 hours) [30] and not used as a means of isolation.<br>- Thus, we assumed the proportion hospitalized was equivalent to the ageadjusted severe disease proportion calculated for the population and applied this to incident infections from the model simulations [21]. We assumed 32% of severe cases would require intensive care [24]. |

| |
|---|
| - **Doubling Rate (Days):** Camp-population Scenario: very fast doubling rate (every 2.3 days) due to the very dense and vulnerable population of IDPs within the camps<br>- This study used a medium **clinical attack rate** for all scenarios, which means 20% of the population will be infected and become symptomatic<br>- In all scenarios, we modelled based on a t**argeted testing strategy**<br>- **Timeframe:** For each scenario, we observed COVID-19 estimated data in terms of cumulative cases, surveillance and lab needs, severe cases, critical cases and expected case fatality rate for 8 weeks from the first diagnosed case<br>- **Case fatality rate:** We use the global average CFR, estimated at 5.9 % based on available data on 11 April20, to project deaths<br>- **Severe and critical cases:** Said differently, about 33% of the severe cases are projected to be critical. We use theseestimates, in all the three scenarios, to generate numbers of predicted severe and critical cases.<br>- **Defining critical time-point in health system capacity:** The researchers considered that the health system capacity reaches critical time-points when number of severe and critical COVID-19 patients requiring hospitalization reaches 50% of number of all available ward and ICU beds, respectively. Considering the total number of ward beds (2,148) and ICU beds (240), this translates to 1,100 ward beds and 120 ICU beds |
| None explicitly stated. |

| |
|---|
| None explicitly stated. |
| None explicitly stated. |

| |
|---|
| None explicitly stated. |
| None explicitly stated. |

| |
|---|
| None explicitly stated. |
| None explicitly stated. |

| |
|---|
| None explicitly stated. |
| None explicitly stated. |
| None explicitly stated. |

| |
|---|
| None explicitly stated. |
| None explicitly stated. |

| Research Questions |
| --- |
| None explicitly stated. |
| None explicitly stated. |

1. Predict the numbers of cases, including severe and critical ones, and deaths.
2. Identify critical time points when the health system capacity is overwhelmed due to COVID-19.

| |
|---|
| None explicitly stated. |

| |
|---|
| None explicitly stated. |
| None explicitly stated. |

| |
|---|
| None explicitly stated. |
| None explicitly stated. |

| |
|---|
| None explicitly stated. |
| None explicitly stated. |

| |
|---|
| None explicitly stated. |
| None explicitly stated. |
| None explicitly stated. |

| |
|---|
| None explicitly stated. |
| None explicitly stated. |

| Research Method(s) |
| --- |
| we implemented a simple stochastic susceptibleexposed-infected-recovered (SEIR) model to estimate the rate of COVID-19 transmission within 111 ICE detention facilities and then examined impacts on regional hospital intensive care unit (ICU) capacity. Models considered three scenarios of transmission (optimistic, moderate, pessimistic) over 30-, 60-, and 90-day time horizons across a range of facility sizes |
| We used a stochastic Susceptible Exposed Infectious Recovered (SEIR) mathematical model to simulate transmission in this population |

| |
|---|
| using the WHO COVD-19 Essential Supplies Forecasting Tool (COVID-ESFT) and data from the Health Information System Unit on population and health facility capacity and utilization in northwest Syria, we generate predicted numbers of cases, deaths and health care needs according to three scenarios. |
| This was a secondary analysis of epidemiologic surveillance data of Mexico from January 1 to May 3 2020 […] We compared the demographic and clinical characteristics, risk conditions, and epidemic curves for migrants and non-migrants. Also, we estimated the cumulative incidence for nonmigrants, and for migrants in two scenarios defined by different estimations of their population size. |

We used two brief screening instruments i.e. Patient Health Questionnaire-2 (PHQ-2) (Kroenke et al., 2003) and Generalized Anxiety Disorder- 2 (GAD-2) (Skapinakis, 2007) to assess depression and anxiety respectively. [...] Perceived stress scale-4 (PSS-4) was used to assess perceived stress which has been reported to be the most useful and feasible in the situations where a short questionnaire is required such as telephonic interview (Lee, 2012). Additionally, a self-designed questionnaire was used to assess the emotional and behavioural response to the lockdown. All the participants were administered these questionnaires by a trained Clinical Psychologist. The data collected were analyzed using SPSS 20.0 version, and descriptive statistics were applied.

The number of daily new cases of COVID-19 in March, April and early May 2020 in Singapore; the cause of a surge of cases in April 2020; the national response to the huge increase in cases; and regulations on migrant worker accommodation were examined. Information was gathered from daily reports provided by the Ministry of Health, Singapore. The principal legislation relating to the housing of migrant workers in dormitories was examined from the Singapore Statues online website. Additional information was gathered from a Ministerial statement given in a Singapore Parliament sitting on 4 May 2020.

| |
|---|
| Continuous variables, if not otherwise specified, are presented as medians (IQR-Interquartile ranges). Categorical variables were described with absolute and relative (percentage) frequencies. Chi-squared and Fisher exact tests were used to compare qualitative variables when appropriate. A two-tailed p-value less than 0.05 was considered statistically significant |
| Population health indicators, incidence, confirmations, and lethality are presented in the following tables as frequency measures; median and interquartile range (25th and 75th percentiles) are shown in Table 1 along with W Shapiro tests for normal distribution, where p > 0.05 indicated normality. From case by case data, we present proportions, standard error, and confidence intervals (all shown as percentages) for age groups, gender, migratory status, confirmation, hospitalization, and survival outcomes. Our results are divided by confirmation, hospitalization, and survival outcome groups. Poisson regression models were used to assess state-cluster indicators and incidence, confirmation, or lethality as dependent variables, after adjusting by population age composition and gender, where the incidence rate ratio (IRR) was the measure of association (reference value of IRR = 1.00). In the case by case analysis, logistic regression for binary dependent variables was used to establish associations between age, gender, and place of residence of patients with the confirmation status, hospitalization, and fatal outcomes; odds ratio was the measure of association, where OR = 1.00 was the reference value. All statistical tests were developed using Stata 14 |

| |
|---|
| Statistical procedures were performed using STATA 11.1 software (StataCorp LLC, USA). Percentage differences were tested using Pearson’s chi-square or Fisher’s exact tests when appropriate. Means of quantitative data were compared using Student's t-test. A p value <0.05 was<br>considered statistically significant. A separate multivariate logistical regression analysis was used to identify independent risk factors for SARS-CoV-2 carriage prevalence among all individuals and in selected groups (when positive cases were found) |
| An anonymized set of data was extracted on April 25 from the electronic patient record containing the following parameters: age, sex, nationality, country of birth, city of residence, date of COVID-19 diagnosis (clinical), date and results of SARS-CoV2 PCR, clinical evolution / outcomes (hospital admission, critical care admission, hospital discharge, length of hospital stay, and in-hospital death). For the present study, patients whose residence was outside the city of Alcorcón were excluded.<br>Statistics. Results are described as means (± standard deviation), medians (interquartile range) or percentages as appropriate. Quantitative variables were compared by the student’s t test, ANOVA, or U Mann-Whitney ‘s test, as appropriate. Qualitative variables were compared by the Chi2 test or theFisher’s exact test as appropriate.<br>Aggregated data by world zone (or country), sex and age group (<30, 30-39, 40-49, 50-59, 60-69, 70-79, 80-89 and >=90) were analyzed. Incidence rates and 95% exact Poisson confidence intervals were calculated by zone. Multivariate negative binomial regression model with robust variance was used to estimate the incidence rate by world zone adjusted for sex and age. These models are appropriate to analyse over-dispersed count data 14. Statistical significance was assumed for p values <0·05. All statistical analyses were conducted with Stata 14 (StataCorp LLC, Texas, USA). The protocol received the approval of the local Institutional Review Board / CEIm (Medicines Research Ethical Committee). |

| |
|---|
| Clinical parameters were stratified to determine sensitivity, specificity, positive predictive value (PPV) and negative predictive value (NPV) according to PCR results. Receiver operating characteristic (ROC) curves were drawn to determine discrimination threshold |
| an anonymous, online survey based on a self-designed, structured questionnaire was conducted.<br>The survey data collected anonymously do not require approval by the local bioethical committee in Poland. Potential respondents were informed in the invitation message about the general subject matter of the survey, its voluntary and fully anonymous character (including no IP tracking). The students were asked to provide answers to questions exploring whether they have faced any prejudice during their stay in Poland (on the street, at restaurants, during shopping, at health service units, and at university) since the outbreak began in China and if such was the case, to briefly describe their experiences and to indicate how much it affected them on 0–5 Likert scale (0 – not at all; 5 – very much). Considering that wearing the face mask could play a potential role in COVID-19-related prejudice, we have identified the frequency and primary cause of their use. Demographic data on gender, age, and time of stay in Poznan´ was also collected. |
| Questionnaire was filled out independently from the participants of several places in Hubei especially in Wuhan. |

| |
|---|
| We administered a phone-based survey to a sample of 1,255 households between April 11 and 17, 2020 to assess the health status, health behaviors, and livelihoods of households across Cox's Bazar. Of those, 909 were reachable by phone and 899 consented to be surveyed. The sample for this study was taken from the Cox's Bazar Panel Survey (CBPS), a longitudinal study tracking 5,020 households across Cox's Bazar that is divided almost equally between refugee camps (n=2,493) and host communities (n=2,527). The Primary Sampling Units (PSUs) in host communities are mauzas, the lowest administrative unit in Bangladesh. We stratified mauzas into areas within 15 kilometers from camps and areas farther away from camps. The PSU for refugee communities were camp blocks, as defined by the International Organization for Migration Needs and Population Monitoring Round 12 (NPM12). |
| A self-report questionnaire was designed to survey peritraumatic psychological distress during the epidemic. |

| Study Population | |
|---|---|
| Demographics | |
| **Definition of migrant group (if any)** | **Migrant group (asylumseekers, refugees, IDPs, undocumented migrants)** |
| ICE detainees | - |
| Rohingya refugees from Myanmar | refugees |

| | |
|---|---|
| internally displaced persons (IDPs) in camps and tented settlements | IDPs |
| The population of interest in this study are persons who are part of mixed migrant flows (economic migrants and asylum seekers), of non-Mexican nationality, who are in Mexico or transiting through the country, many of them with the intention of requesting asylum in the United States | asylum seekers and economic migrants |

| | |
|---|---|
| migrant workers identified by the Government of India, who were living in the shelter house or government authorized buildings | - |
| Migrant workers in Singapore | - |

| | |
|---|---|
| Migrants | - |
| Migration. This includes the totality of national and international migratory movements projected for the year 2020 from CONAPO estimates | - |

| | |
|---|---|
| asylum seekers | asylum seekers |
| Patients were classified according to their country of origin into one the following groups | - |

| | |
|---|---|
| migrant workers residing in a large dormitory in Singapore | - |
| Asian students at Poznan University of Medical Sciences in Poland | - |
| international students living in Hubei province | - |

| | |
|---|---|
| Rohingya refugees | Rohingya refugees |
| migrant workers | - |

| **Other legal categories such as migrant workers or not specifically defined in international law such as international students** | **Country of Study** |
|---|---|
| - | USA |
| - | Bangladesh |

| | |
|---|---|
| - | Northwest Syria |
| - | Mexico |

| | |
|---|---|
| migrant workers | India |
| migrant workers | Singapore |

| | |
|---|---|
| - | - |
| interstate migrants (not further specified) | Mexico |

| | |
|---|---|
| - | France |
| migrants (as in all nationalities different from Spain) | Spain |

| | |
|---|---|
| migrant workers | Singapore |
| international students | Poland |
| international students | China |

| | |
|---|---|
| - | Bangladesh |
| migrant workers | China |

| **Socioeconomic Development of Country/Region** | **Age/Age-groups** |
| --- | --- |
| HIC | majority of immigrants detained by ICE are between 26 and 35 years old, where the median age is 30 |
| LIC | Median age: 16 years |

| | |
|---|---|
| LIC | not explicitly stated. |
| Upper MIC | (0-17y, 18-49y, and +50y) |

| | |
|---|---|
| low MIC | mean age of 32.7 (SD: 10.1) years |
| HIC | not stated. |

| | |
|---|---|
| - | Migrants were younger than natives: in cohort A the median (IQR) age was 40 (27-49) VS. 66 (46-70) years, whereas in cohort B 37 (27-46) VS. 48 (47-60) years. |
| Upper MIC | (?) three groups (<15, 15–65 and >65)<br>COVID-19 cases in the studied period were 55.76% female, and the mean age was 43.4 years (±0.49). An amount of 2.8% (4.97) of patients were aged <15, and 12.2% (2159) were aged 65 and over; by 21 April, 17,763 cases had been registered in the 32 states. Age distribution did not significantly differ between states or by gender (male 43.8 years old, female 44.8 years old). Description of state-cluster indicators and COVID-19 cases per state-cluster are presented in Table not sure if this is also applicable for migrant group |

| | |
|---|---|
| HIC | range: 0-67 years<br>Mean (SD): 21.6 (+-13.6) |
| HIC | age group (<30, 30-39, 40-49, 50-59, 60-69, 70-79, 80-89 and >=90)<br>- 52 years medan age |

| | |
|---|---|
| HIC | mean age: 33 years |
| HIC | mean ± SD age 23.8 ± 3.8; |
| upper MIC | average age: 30 years (SD=9.26) |

| | |
|---|---|
| LIC | - 15-24 (24%)<br>- 25-34 (30%)<br>- 35-44 (11.7%)<br>- 45+ (14.7%) |
| upper MIC | not explicitly stated but from results one can say:<br>(only general population)<br>- under 18<br>- between 18-30<br>- above 60 |

| country of origin | Sex |
| --- | --- |
| Country of origin for detainees included in this analysis was not available; | we obtained facility-level sex data: There were 577 women held in two female-only facilities; 15,560 individuals were held in 43 maleonly facilities; and 26,208 detainees were in facilities that held both females and males |
| Myanmar | not accounted for |

| | |
|---|---|
| Syria | not explicitly stated. |
| Central America, the Caribbean, Venezuela and Africa | 43,2% women |

| | |
|---|---|
| Not explicilty stated. | all male |
| South Asia | mainly male |

| | |
|---|---|
| - | not explicitly stated. |
| not explicitly stated. | 55.76% female |

| | |
|---|---|
| europe: 15.6%<br>Africa: 41.6%<br>Asia: 42.9% | 35.1% Female (64.9% male) |
| country other than spain (table 1)<br>Regions: European region (including switzerland and Norway)<br>Eastern Europeand Russia<br>Asia<br>Northern Africa<br>Sub-saharan Africa<br>Latin America<br>Carbbean<br>Miscellanea (US, Canada, Australia, NZ) | 52.5% male |

| | |
|---|---|
| Not stated. | Not stated. |
| Asia, mostly from Taiwan (75,3%) | 49 females, 36 males |
| Not explicitly stated. | 57.1% male (42.9% female) |

| | |
|---|---|
| Not explicitly stated. | male: 60.2% (39.8% female) |
| Not stated. | only for general population (male: 35.27%) female (64.73%) |

Further relevant information

| SES | Further relevant characteristics |
|---|---|
| - | however, a 2019 report by the U.S. Government Accountability Office found that the majority of detentions from 2015 to 2018 were males from Mexico, Guatemala, El Salvador, and Honduras, without a previous arrest or conviction record |
| - | (Abstract:Background:)with 600,000 concentrated in the Kutupalong-Balukhali Expansion Site (mean age, 21 years; standard deviation [SD], 18 years; 52% female). |

| | |
|---|---|
| - | Up to 41% of the adult Syrian population has a non-communicable disease e.g. hypertension, diabetes, cancer; . Smoking prevalence among Syrian adults (particularly men) is among the highest in the Middle East and is associated with more severe disease5 . Malnutrition (micro and macronutrient deficiency) is high given food insecurity in the area6. |
| - | - |

| | |
|---|---|
| the mean a number of years of education being 2.4 (SD: 1.7) years. The mean income of the participants before the lockdown was 8280 Indian rupees. | Majority of the participants were married (69.4 %). |
| employed in the construction, marine and other low wage sectors | low-skilled migrant workers living in dormitories scattered throughout Singapore |

| | |
|---|---|
| - | Migrants had fewer co-morbidities than natives; in particular, 23/43 (53.5%) migrants had no co-morbidities versus 5/26 (19.2%) natives (p-value: 0.005). |
| - | - |

| | |
|---|---|
| - | Residence γ is dedicated to asylum-seekers, including family groups and single individuals |
| - | - |

| | |
|---|---|
| - | - |
| - | All of them have been living in Poznan´ for at least half a year (with a mean ± SD of 2.7 ± 1.4 years). |
| Similarly, the students who were doing bachelor and PhD were more likely to be affected (Odd Ratio = 0.66) than the master students | Of which 368 (73%) were married and 136 (27%) were single |

| | |
|---|---|
| Data from the 2019 CBPS baseline survey shows that refugee households have significantly lower levels of income and assets (Table 1) compared to members of the host community.<br>The poorest households (first quintile) in refugee and host communities hold assets for an average value of 5.5 USD and 224.2 USD ($p < 0.001$), respectively. | Educational level (2019), Household assets value (2019 USD); Household income (2019 USD), Employment (2019-2020); Food insecuritiy (2020); Water, sanitation and crowding (2019); Trauma and Clinical Depression (2019) --> vgl. Tabelle 1 |
| - | - |

| Sample Size(migrant group) |
| --- |
| 42,435 individuals detained. -Thirty-two percent (n = 35) of the included facilities hold only ICE detainees, and 68% (n = 76) are city or county correctional facilities that hold ICE detainees and other individuals who are incarcerated |
| 600.000 |

| |
|---|
| 1.2 Million |
| 74 |

98

As of 6 May 2020, there were **17758** confirmed COVID-19 cases among those living in migrant worker dormitories

43

77

n= 20419

| |
|---|
| n=5977 |
| n=85 |
| 504 |

| |
|---|
| n=367 |
| not stated |

| | Outcomes, Exposures and Co-Variables |
|---|---|
| | Housing Exposures/Predictors |
| **Sampling strategy** | **Individual-level** |
| ICE detainee population data were downloaded directly from the Department of Homeland Security Enforcement and Removal Operations (ERO) website. The average number of detainees between October 1, 2019 and March 2, 2020 was used to approximate the current number of people detained at each detention facility and summed to approximate the total number of people currently detained by ICE. | - |
| using the Rohingya refugees living in the Kutupalong-Balukhali Expansion Site as a case study | - |

| | |
|---|---|
| 1.2 Million | - |
| The information was extracted from a public data base of suspect cases published by the Mexican Ministry of Health, with cutoff date May 3, 2020 (first date of symptom onset January 1, 2020).The data base from the Ministry of Health includes a dichotomous variable coded migrant/nonmigrant. This variable does not indicate migration status (regular vs. irregular), and the information is missing in 99.5% of cases. Therefore, to identify our population of interest, we used the variable "country of nationality", and selected those whose nationality corresponded to one of the main sending countries or regions of mixed migrant flows in Mexico: Central America, the Caribbean, Venezuela and Africa (Cobo & Fuerte, 2012; Rodriguez, 2016). | - |

| | |
|---|---|
| The migrants' workers identified by the Government of India, who were living in the shelter house or government authorized buildings, were recruited | - |
| cases taken from different sources | - |

| | |
|---|---|
| Not explicitly stated. | - |
| Data were obtained from open-access epidemiologic data of COVID-19 available from the DGE webpage, including laboratory-confirmed and clinically diagnosed (suspected) patients.<br>Case by case data were obtained from the open-access DGE dataset for COVID-19 during the imported case and community transmission phases, including all registries up to 21 April 2020. | - |

| | |
|---|---|
| Based on the preliminary information that some homeless persons from these two shelters presented with COVID-19 symptoms, we organised a screening campaign in collaboration with the staff in charge of these shelters. We subsequently received other requests for screening from several accommodation centres specialising in housing vulnerable people.<br>Participants were encouraged by the management staff of the facilities to get tested and were then recruited on a voluntary basis. | family appartments, capacity of 50, private bathrooms and kitchen, no open spaces. |
| Case was defined as a patient with a COVID-19 diagnosis at Hospital Universitario Fundación Alcorcón confirmed by Polymerase Chain Reaction (PCR) for SARS-CoV2 for adults residing in Alcorcón. Incident cases of COVID-19 were obtained from the Electronic Patient Record (Selene ©) that is used for all medical and administrative interactions with patients. COVID-19 clinical diagnosis was established by the attending clinicians at the Emergency Room according to European Center for Disease Control and Prevention (ECDCP)-World Health Organization Criteria (WHO) 13. | - |

| | |
|---|---|
| With the high number of COVID-19 cases arising from dormitory A, it was gazetted by law as an isolation area on April 5th [2]. We describe results of the outbreak investigation and clinical evaluation in dormitory A. | - |
| The selected group included Asian students at Poznan University of Medical Sciences in Poland who were directly invited to complete the survey via an e-mail message. All surveyed students were living in the city of Poznan´ (Greater Poland Voivodeship). | - |
| We approached students through official WeChat groups, which were already developed by the universities for international students. | - |

| | |
|---|---|
| The baseline survey of the CBPS was collected between April and July 2019. In each household, we administered a household-level questionnaire covering a nu mber of topics including the value of assets held by the household and income from different sources. I n addition, we randomly selected two adults aged 15 or older for detailed interviews covering a wide ran ge of topics, including detailed questions on labor market outcomes and trauma and mental health. For t he present survey, we asked to speak with at least one of the two randomly selected adults. We were abl e to interview 704 out of the 909 randomly selected adults that were interviewed in 2019. In household s where none of the adults interviewed in 2019 were available to be interviewed, we administered the ques tionnaire to another adult member. | As expected, housing conditions that favor community transmission are more often observed in camps (Table 1). Only 1.2% of households in camps, but 55.3% of those in host communities, have a private toilet p<0.001) and as many as 31.3% of households in camps share a toilet with more than 25 people, compared to 0% in the host community (p<0·001). Sharing a water source with a large number of users is also commonplace in camps, where 62.1% report sharing water facilities with more than 25 users, whereas only 6.6% of the host community do so (p<0·001). |
| Leveraging the Siuvo Intelligent Psychological Assessment Platform, we presented QR codes of the questionnaire online openly accessible to the general public nationwide. | - |

| Contextual-level |
| --- |
| Only facilities with more than 25 people |
| 23 congested settlements, With over 46,000 persons per square kilometer |

-

-

| |
|---|
| living in shelter house or in government authorized buildings |
| - |

| |
|---|
| - |
| - |

| |
|---|
| 102 residents and 19 Employees; 78.5% tested (out of all 121 people) an 75.5% tested residents. 17 days between first day of lockdown and screening (days) |
| - |

| |
|---|
| Dormitory A is a large privately owned dormitory that can accommodate 13,000 inhabitants, in rooms of 12 residents across 13 blocks. |
| - |
| - |

| |
|---|
| - |
| - |

| Non-pharmaceutical intervetions and policies/strategies implemented | |
|---|---|
| **Individual-level** | **Contextual-level** |
| - | - |
| - | - |

| | |
|---|---|
| - | - |
| - | - |

| | |
|---|---|
| - | - |
| Massive logistic arrangements have been made to provide housing facilities that will allow for appropriate social distancing. These included vacant public housing flats, military camps, exhibition centres and even floating hotels. Preparations were put in place for food delivery, hygiene maintenance, monitoring and enforcement of quarantine, WiFi for workers to communicate with family and for entertainment and distribution of 'care packs' that contained reusable masks, hand sanitisers and thermometers. | Among the sweeping measures implemented were extensive testing of dormitory workers, segregation of healthy and infected workers, observation for fever and symptoms several times a day, and setting up of dormitory on-site healthcare facilities. The local community also helped in the response. Within days, websites that offered English to Bengali (tinyurl.com/covidbengali) and English to Tamil translations (http://better.sg/migrantw orkertranslations) to medical care teams were developed. This helped overcome the language barrier and allowed non-Bengali and non-Tamil-speaking healthcare workers to conduct an initial consultation without an interpreter. The websites also enabled medical personnel to contact a group of volunteer interpreters directly. About 3000 healthcare professionals signed up to the SG Healthcare Corps since its launch in April to marshal volunteers. Many were deployed to assist in the management of the dormitory infections. |

| | |
|---|---|
| - | - |
| - | - |

| | |
|---|---|
| - | All residents of homeless shelters were placed under strict lockdown since 17 March, in line with the whole French population (=C0), allowing all homeless people to stay in the shelter 24 hours a day |
| - | - |

| | |
|---|---|
| - | - |
| **wearing face masks:** Wearing the face mask was relatively common within the studied group, with 43.5% of students declaring that they wear them regularly, particularly in December, January and February | - |
| Many respondents obeyed precautionary measures to avoid COVID-19. They reduced contact with others (78.96%), decreased visits to the affected areas (71.03%), increased the frequency of washing hands (95.83%), and took more care of their room ventilation (88.88%). While attending public places also declined (94.84%) (Fig. 1d). | - |

| | |
|---|---|
| - | - |
| - | Three major events during the COVID-19 epidemic may have caused public panic: (1) the official confirmation of human-to- human transmission of COVID-19 on 20 January; (2) the strict quarantine of Wuhan on 22 January and (3) WHO's announcement of PHEIC on 31 January. This study began on 31 January. |

| SARS-CoV-2 / COVID-19 related outcomes | |
|---|---|
| **Outcome of interest (SARS-CoV-2 / COVID-19)** | **Instruments to capture SARS-CoV-2 / COVID-19 related exposure** |
| COVID-19 Infections | modeling study dependent on the reproductive number (R0) |
| Transmission scenario of COVID-19 | modeling study dependent on the reproductive number (R0) |

| | |
|---|---|
| COVID-19 Cases | modeling study (see assumptions) |
| COVID-19 cases<br><br>COVID-19 incidence | According to information on the downloading site, these are preliminary data (not yet validated) collected by the Epidemiological Surveillance System for Viral Respiratory Disease (previously Epidemiological Surveillance System for Influenza -SISVEFLU-), which are captured by 475 Viral Respiratory Disease Monitoring Units (USMER), throughout the country. |

| | |
|---|---|
| - | - |
| SARS-CoV-2 Infection | cases of COVID-19 are confirmed by a positive finding of a nasal swab which undergoes PCR testing for SARS-CoV-2 |

| |
|---|
| Descriptive only |
| COVID-19 Incidence |

| | |
|---|---|
| COVID-19 positive cases | Participants were encouraged by the management staff of the facilities to get tested and were then recruited on a voluntary basis. They were systematically asked to provide basic demographic information (sex, age, country of origin), chronic conditions, and any respiratory symptoms or fever in the two weeks prior to sampling. Body temperature was measured using a forehead infrared thermometer. Nasal samples were systematically collected on transport media using Sigma Transwabs (Medical Wire, Corsham, United Kingdom). For self-sampling, participants were invited to insert the swab into their nostrils (about 2 cm). If individuals were unable to perform selfsampling, trained investigators carried out the sampling. Specimens were immediately processed for SARS-CoV-2 PCR testing. Homeless peoples' pets were also tested [illegible] |
| 1.) COVID-19 Incidence rate<br><br>2.) Relative risk ratio for COVID-19 | For molecular diagnosis of SARS-CoV-2 infection, nasopharyngeal swabs, sputum, or bronchopulmonary aspirates were processed by automatized extraction using the MagNaPureLc instrument (Roche Applied Science, Mannheim, Germany) and real time reverse-transcription PCR using the SARS-Cov-2 nucleic acid detection Viasure kit (CerTestBiotec S·L·), following the manufacturer's instructions. For this rRT-PCR, we used Bio-Rad CFX96™ Real-Time PCR Detection System |

| | |
|---|---|
| COVID-19 positive cases | Nasopharyngeal specimens were sent for polymerase chain reaction (PCR) testing for severe acute respiratory syndrome coronavirus-2. |
| Preconceptions related to COVID-19 | 5 Point-Likert scale |
| - | - |

| | |
|---|---|
| Prevalence of COVID-19 Symptoms | We administered a checklist of sy mptoms based on the WHO and C DC guidelines. We used the three most common sym ptoms featured on the WHO dedic ated COVID-19 website on April 27 , 2020 to produce our preferred measure of COVID-19 risk: having at least one of the symptoms (feve r, dry cough, and fatigue or tiredness). |
| - | - |

| **Measures of frequency/ associations for each outcome (if applicable)** | **CI lower** |
|---|---|
| different time horizons: 30, 60 and 90 days; and different R(0) scenarios | with R(0)=2.5<br>50 (size)<br>100<br>500<br>1000<br>with R(0)=3.5<br>50<br>100<br>500<br>1000 |
| scenario: low, moderate, high and different time horizons: 1 month, 3 months and 12 months | In all scenarios, we observed relatively slow growth during the beginning of the simulated outbreaks (Fig 1), with limited numbers of people infected and few, if any, hospitalizations and deaths during the first month after the introduction of one infectious case (Table 3). However, this quickly changed once sufficient infections were in the population, with cases rapidly increasing and the outbreaks culminating within the year (Table 3, Fig 1). By the time the first hospitalization occurs, we expect 50 (95% PI, 1–197) individuals to be infected in the population under the low scenario, assuming homogeneous probability of infection by age. This increases to 72 (95% PI, 2–289) and 141 (95% PI, 3–502) in the moderate and high scenarios, respectively |

| | |
|---|---|
| Within the first six weeks, the COVID-ESFT projects a total cumulative case load of 240,000, equating 20% of the total population of the camps (1,200,000) | - |
| COVID-19 cases: 13,9% positive cases in migrants compared to 28% positive cases in people of Mexican nationality.<br><br>COVID-19 Incidence: The estimated cumulative incidence for migrants and non-migrants is illustrated in Figure 3. The graph shows that, in either of the two scenarios, the number of suspect cases per 100,000 would be higher among migrants. The cumulative incidence ratios comparing migrants with nonmigrants are 6.12 (CI95% 4.75, 7.77) for the first scenario, and 1.49 (CI95% 1.15, 1.89) for the second scenario | Cummulative incidence ratio scenario 1: 4.75<br><br>Cummulative incidence ratio scenario 2: 1.15 |

| | |
|---|---|
| - | - |
| The majority of the new cases occurred among low-skilled migrant workers living in dormitories scattered throughout Singapore. As of 6 May 2020, there were 17758 confirmed COVID-19 cases among those living in migrant worker dormitories (88% of 20198 nationally confirmed cases).6 A single dormitory housing approximately 13000 workers had 2526 confirmed cases, accounting for 12.5% of all cases in the country | - |

| | |
|---|---|
| In the Poisson regression model (Table 2), a significantly higher incidence rate ratio was observed with a higher migration rate (IRR = 6.43:1),<br><br>*(Migrantion showed 6,43 times the rate of coronavirus infection than the indigenous ethnicity)* | - |

| | |
|---|---|
| One employee was tested positive, none of the tested residence was tested positive | - |
| 1.) By origin, the global accumulated COVID-A-PCR+ incidence in Spaniards was 6·50 cases per 1000 inhabitants as compared to 8·82 per 1000 for non-Spaniards.<br>The global cumulative incidence increased dramatically with age for<br>both Spaniards and migrants.<br><br>2.) The adjusted relative risks for COVID-PCR+ among migrants from Asia, European Union and Eastern Europe did not significantly differ from Spain (table 3). In contrast, the relative risk for Latin-America migrants was about 7-fold higher than that for Spaniards (RR 6·92; 95% CI 4·49- 10·67, p <0·001). In addition, the adjusted risk was also increased for Sub-Saharan Africa (RR 3·66; 95% 1·42-9·41, p=0·007) as well as Caribbean (RR 6·35 95% CI 3·83-10·55, p<0·001) in a highly clinical and statistically | Spaniards: 4·49- 10·67<br>Sub-Saharan Africa: 1·42<br>Caribbean: 3·83 |

| | |
|---|---|
| n all, 1832 out of 5977 foreign workers were symptomatic. Of these, 1264 (69%) were found to be positive for COVID-19. | - |
| Last but not least, nearly one quarter (24.7%) of Asian students have faced preconceptions related to COVID-19 at the university where they study.<br>Again, these situations had a negative effect on most of the surveyed students - a score of 5 was most frequently selected (median; IQR: 4; 3–5) | - |
| - | - |

| | |
|---|---|
| Respondents in refugee camps are almost twice as likely as hosts to report having had fever<br>(13·9% and 6·6% respectively, p=0·001) and dry cough (9,5% and 5,4% respectively, p=0,037) in the previous week (Table 2).<br>Furthermore, camp residents are more like ly to show<br>at least one of the three most common sy mptoms of COVID-19 (18·1% and 11·0% res pectively, p < 0·001)<br>[...] Taken together, these results suggest that the observed differences in self-reported health are<br>concentrated specifically in COVID-19 symptoms. | - |
| - | - |

| CI upper | Standard Error |
| --- | --- |
| 95%CI at 30 days:<br>25 (7,36)<br>36 (7,62)<br>60 (12,128)<br>494 (491,495)<br>95%CI at 30 days:<br>33 (16,42)<br>59 (24,80)<br>136 (36,232)<br>163 (40,309) | - |
| | - |

| | |
|---|---|
| - | - |
| 7.77<br><br>1.89 | - |

| | |
|---|---|
| - | - |
| - | - |

| | |
|---|---|
| - | - |

| | |
|---|---|
| - | - |
| Spaniards: 10·67<br>Sub-Saharan Africa: 9·41<br>Caribbean: 10·55 | - |

| | |
|---|---|
| - | - |
| - | - |
| - | - |

| | |
|---|---|
| - | - |
| - | - |

| Other health-related outcomes reported | |
|---|---|
| **Physical health** | **Mental health** |
| - | - |
| - | - |

| | |
|---|---|
| - | - |
| Risk conditions:<br>Hypertension: 8.1% (migrants); 20,6% (mexicans)<br>Diabetes: 2,7% (migrants); 15.2% (mexicans)<br>COPD: 1.4% (migrants); 2.3%(mexicans)<br>Asthma: 8.1% (migrants); 5.5%(mexicans)<br>Obesity: 4.1 % (migrants); 16.4% (mexicans)<br>Immunosupression: 2.7% (migrants); 2.7% (mexicans)<br>Pregnancy: 9.4% (migrants); 2.6% (mexicans) | - |

| | |
|---|---|
| - | About three fourth of the participants (73.5 %) were found to be screen positive for depression on the PHQ-2 and about half of the participant (50 %) were found to be screen positive for anxiety on the GAD-2 (Table 2). On PSS-4, the mean score on the PSS was 7.1 (2.3). About one-fifth of the participants screened positive for depression only. Nearly half (51 %) of participants screened positive for both anxiety and depression. Overall, about three-fourth (73.5 %) screened positive for at least one psychiatric morbidity (Table 1). |
| - | - |

| | |
|---|---|
| Migrants had fewer co-morbidities than natives; in particular, 23/43 (53.5%) migrants had no co-morbidities versus 5/26 (19.2%) natives (p-value: 0.005) | - |
| - | - |

| | |
|---|---|
| Presence of respiratory symptom (cough, rhinirrhoea, Dyspnoea, sore throat) and fever:<br>At least one symptom: 6.5% | - |
| - | - |

| | |
|---|---|
| COVID-19-positive and -negative groups had similar median age (33 years, P ¼ 0.71) and heart rate (108 vs 107 bpm, P ¼ 0.17). COVID-19-positive foreign workers had slightly higher median temperatures (37.8 vs 37.7, P < 0.01). | - |
| - | - |
| - | **socio-psychological impact** of COVID-19 on their daily life<br>Approximately 71.03% used to talk about virus in their regular talks, while 49.60% were unhappy due to this pandemic condition. At one point, 50.19% had their concern about virus transmission, 48.21% were much worried about their family safety and 23.21% felt helpless (Fig. 1c). |

| | |
|---|---|
| - | Surprisingly, given the link between psychological stress and immunity,14-16 we find that lifetime trauma and depression severity are not significantly correlated with COVID-19 symptoms |
| - | level of distress |

| Social well-being |
| --- |
| - |
| - |

-

-

On the self-designed questionnaire, about two-thirds (63.3 %) of the participants reported the markedly increased in the loneliness. More than half of the participants said a significant increase in tension (58.2 %), frustration (58.2 %), low mood (55.1 %), irritability (51.0 %), and fear of death (51.0 %). The other more common responses were fear (41.8 %) and social isolation (31.6 %). There was a marked reduction in the social connectedness (48 %) and sleep (44.9 %) among the participants (Table 2)

Twenty-four dormitories have been declared as isolation areas, with residents quarantined for 14 days.

| |
|---|
| - |
| - |

-

-

-

**Prejudice**: We have found that 61.2% of the surveyed students have experienced prejudice in Poland related to the current coronavirus epidemic, and it was more frequently witnessed by those wearing face masks than those who do not (71.2% vs 28.2%). The prejudice most commonly encountered, by 47.1%, in public transportation and on the street.

**- prejudice in public transport:** using a Likert scale, a score of 5 was the most frequently selected (median; IQR: 3; 2–5)

**- prejudice in restaurants or shopping:** These reactions had an obvious negative effect on Asian students, since 5 was the most frequently selected score when evaluating them on the Likert scale (median; IQR: 3; 2–5).

**- prejudice in health services:** These reactions had an obvious negative effect on Asian students, since 5 was the most frequently selected score when evaluating them on the Likert scale (median; IQR: 3; 2–5).

**In another two specific questions about to stay in China and the policies by Chinese government**, 92.85% students were satisfied with the steps took by government and 58.53% preferred to stay here in China rather than to go their home countries

**Knowledge about COVID-19 and health behaviors**. A survey module administered to a subsample of respondents revealed that trusted sources of advice on COVID-19 prevention vary greatly across refugees and hosts, but information provided by friends and acquaintances is important for both (58·8% and 62·9% of respondents respectively, p=0·437) (Table 4). Among refugees, NGOs are also trusted sources (53·5%), followed by informational campaigns on the street (41·6%) and local leaders (e.g., block majhees). Among hosts, newspapers, radio, and TV are the most trusted sources of information (81·4%), and social media is cited by many (51·7%)

- Also information about importance of good respiratory and household hygiene practices (vast majority of respondents are cognizant of the importance)

-Lastly, we find some evidence that fear is breeding stigma in some communities (Table 5). Nearly one-third of refugees and hosts (30·9% and 35·1% respectively, p=0·406) report that suspected carriers of COVID-19 were prevented from receiving treatment in their community

---

Results also indicated that as time passes, distress levels among the public have been significantly descending, with the lowest distress level during the Lantern Festival (8 February).

| Instruments used to capture health conditions |
| --- |
| - |
| - |

-

-

| |
|---|
| PHQ2, GAD2, PSS-4, Additionally, a self-designed questionnaire was used to assess the emotional and behavioural response to the lockdown. |
| - |

-

-

-

-

| |
|---|
| Oxygen saturations and heart rate were measured using portable pulse oximeters. |
| - |
| - |

| |
|---|
| We measured<br>lifetime trauma using the Harvard Trauma Questionnaire (HTQ)8<br>and depressive mood using the 9-item<br>version of the Patient Health Questionnaire (PHQ-9).9<br>We used a cut-off point of PHQ-9 equal to or above<br>10 as a screener for depression.1 |
| The questionnaire incorporated relevant diagnostic guidelines for specific phobias and stress disorders specified in the International Classification of Diseases, 11th Revision and expert opinions from psychiatrists. In addition to demographic data (ie, province, gender, age, education and occupation), the COVID-19 Peritraumatic Distress Index (CPDI) inquired about the frequency of anxiety, depression, specific phobias, cognitive change, avoidance and compulsive behaviour, physical symptoms and loss of social functioning in the past week, ranging from 0 to 100. A score between 28 and 51 indicates mild to moderate distress. A score ≥52 indicates severe distress. |

| hospitalisations |
| --- |
| Under the pessimistic scenario of R0 = 7, and imagining synchronized epidemics cross all facilities, a median of 5145 detainees system-wide will become infected and require hospitalization within the first 30 days, growing to an aggregate of 6391 patients by day 60 and 6408 patients by day 90. Averaging across the total ICE system, this represents 15.1% of all detainees. Concretely, in a facility housing 1000 detainees, the estimated median number of people infected and requiring hospitalization by day 30 ranges from 10 (R0 = 2.5) to 26 (R0 = 3.5) and 116 (R0 = 7) and by day 90 ranges from 114 (R0 = 2.5) to 148 (R0 = 3.5) and 157 (R0 = 7).<br>(median number 95%CI), 90 days size:<br>R(0)=2.5; R(0)=3.5; R(0)=7<br>50: 6; 7; 7<br>100: 13; 15; 16<br>500: 58; 70; 75<br>1000: 114; 148; 157 |
| When the first hospitalization occurs, we expect the virus to have been circulating in this population for an average of 38, 30, and 23 days under the low, moderate, and high transmission scenarios, respectively. Adjusted for the age distribution in the Kutupalong-Balukhali Expansion Site, we estimated that 4.8% (95% PI, 0.3%–15%) of infections in this population would result in severe disease and hospitalization. The maximum daily hospitalization capacity needed ranges between 5,210 (95% PI, 3,120–8,090) in the low transmission scenario and 15,450 (95% PI, 9,500–23,990) beds in the high (Table 2). Under the low transmission scenario, hospitalization needs exceeded the hospitalization capacity of 340 beds after 136 days (95% PI, 96–196 days), while in the high transmission scenario, this occurred after only 55 days (95% PI, 42–77 days;(Table 2, Fig 2)<br>In the high transmission scenario, the maximum number of daily hospitalizations is reached on day 80, compared to day 117 and day 190 on average in the low and moderate scenarios, respectively (Fig 1). Current hospital bed surge capacity (630) as reported from the site may almost double the number of available beds, which would delay overwhelming the capacity by 3–10 days on average |

| |
|---|
| - |
| 10,8% (migrants); 27.7% (mexicans) |

-

-

| |
|---|
| - |
| Incidence Rate ratio: 0.65 for Migration in comparison to indigenous ethnicity (1.01)<br><br>Hospitalizations varied among state-clusters, migration and availability of clinics and hospitals were inversely associated, and urbanization and population affiliated to public health institutions showed higher IRR for hospitalization.<br><br>Table3<br>COVID-19 cases by laboratory confirmation status, hospitalization, and mortality in Mexico between 28 February and 21 April 2020 (n = 17,763)<br>6,49% (95%CI: 6,05; 6,93) (n=5545) of interstate Migrant had to be hospitalized?<br><br>Interstate migrants were less prone to be laboratory-confirmed but showed more susceptibility to hospitalization and death. |

-

-

| |
|---|
| - |
| - |
| - |

-

-

| ICU |
| --- |
| Focusing now on estimating ICU admissions, we estimate that with R0 = 2.5, and again imagining that all epidemics are synchronized, there will be a median of 139 detainees infected who will require ICU admission after 30 days, growing to an aggregate of 475 patients by day 60 and 745 patients by day 90 (approximately 1.8% of the total population). Under the pessimistic scenario of R0 = 7, the corresponding numbers of patients are 782, 971, and 973 on days 30, 60, and 90, respectively. This corresponds to approximately 2.3% of the total population<br>(median number 95%CI), 90 days<br>size: R(0)=2.5; R(0)=3.5; R(0)=7<br>50: 1;1;1<br>100: 2;2;2<br>500: 9;11;11<br>1000: 17;22;24 |
| Within 3 months of successful introduction, we estimated 6 (95% PI, 0–30) cumulative ICU admissions in the low transmission scenario compared to 4,400 (95% PI, 98– 10,100) in the high transmission scenario (Table 3) |

Over 6 weeks, 12,000 cases will be critical, requiring ICU admission. The critical moment for critical cases in this scenario will be by the mid of 4th week when ICU COVID-19 needs exceed 50% of available ICU beds capacity

-

| |
|---|
| - |
| - |

-

-

| |
|---|
| - |
| - |
| - |

-

-

| mortality | Other health-related outcomes (e.g. ventilation ..) |
| --- | --- |
| - | - |
| - | - |

| | |
|---|---|
| The COVID-ESFT predicts 14,328 deaths by the end of week 4. However, both the CFR<br>and the number of deaths are likely to be much higher due to lack of the capacity of the health system to deal with the expected load of severe and critical cases.<br>Within the first six weeks: 18,751 deaths | - |
| - | intubated cases: 12.5% (migrants); 6.0% (mexicans)<br><br>Pneumonia: 5.4% (migrants); 19.5% (mexicans) |

| | |
|---|---|
| - | - |
| - | - |

| | |
|---|---|
| Overall, migrants had a lower mortality rate than natives (1/43, 2.3% versus 7/26, 26.9%; p-value: 0.002). | - |
| Incidence Rate ratio: 1.02 for Migration in comparison to indigenous ethnicity (0.82)<br><br>COVID-19 cases by laboratory confirmation status, hospitalization, and mortality in Mexico between 28 February and 21 April 2020 (n = 17,763) 4,75 % (95%CI: 4,06; 5,43) of Interstate Migrants (n=968 confirmed cases) had a fatal outcome **?**<br><br>Interstate migrants were less prone to be laboratory-confirmed but showed more susceptibility to hospitalization and death. | - |

| | |
|---|---|
| - | - |
| - | Severe COVID-19, defined as death, critical care admission or hospital stay longer than 7 days occurred in 63% of the Spaniards as compared to 33% for migrants. Similarly, unadjusted mortality was higher for Spaniards (25% of admissions) than for migrants (6%). |

| | |
|---|---|
| - | PPV was highest for those with **body temperature** 38.5C (89.0%) (Table I, Figure 1) and **oxygen saturations** of of 88.9%. |
| - | - |
| - | - |

| | |
|---|---|
| - | - |
| - | - |

| Measures of frequency/ associations for each outcome (if applicable) | CI lower | CI upper | Standard Error |
|---|---|---|---|
| - | - | - | - |
| - | - | - | - |

| | | | |
|---|---|---|---|
| - | - | - | - |
| - | | | |

| | | | |
|---|---|---|---|
| - | - | - | - |
| - | - | - | - |

| | | | |
|---|---|---|---|
| - | - | - | - |
| Hospitalisation<br><br>Laetality (Mortality) | 6,05<br><br>4,06 | 6,93<br><br>5,43 | 0.22<br><br>0,68 |

| | | | |
|---|---|---|---|
| - | - | - | - |
| - | - | - | - |

| | | | |
|---|---|---|---|
| - | - | - | - |
| - | - | - | - |
| - | - | - | - |

| | | | |
|---|---|---|---|
| **Treatment-seeking behavior:** For those who experienced at least one symptom of any health condition (see Table 2 for a breakdown), pharmacies were the fir st stop for advice and treatment (69·6% an d 42·3% in host communities and camps respectively, p<0·001) (Table 3). Among refugees, health information providers in camps are the second most-common healthcare provider (35·8% visited one to treat their symptoms). | - | - | - |
| It is noteworthy that migrant workers experienced the highest level of distress: mean=31.89, F=1602.501, p<0.001 | - | - | SD: 23.51 |

| Co-variables/Potential confounders considered | |
|---|---|
| **Individual-level** | **Contextual-level** |
| - | - |
| - | - |

| | |
|---|---|
| - | - |
| - | - |

| | |
|---|---|
| The other confounding factors, including a history of depression, substance use, or physical illness, were not taken | The assessment was crosssectional, and the specific ongoing stressors, coping mechanisms, etc. were not evaluated. |
| - | - |

| | |
|---|---|
| - | - |
| - | - |

| | |
|---|---|
| - | - |
| - | - |

| | |
|---|---|
| - | - |
| - | - |
| - | - |

| | |
|---|---|
| - | - |
| - | - |

| |
|---|
| **Instruments used to capture co-variables** |
| - |
| - |

-

-

| |
|---|
| - |
| - |

| |
|---|
| - |
| - |

-

-

| |
| --- |
| - |
| - |
| - |

| |
|---|
| - |
| - |

| Findings and Limitations |
| --- |
| **Main results/ Conclusions (as reported)** |
| To our knowledge, this is the first analysis assessing the impact of COVID-19 on ICE detainees and the wider communities likely to care for detainees. Across the 111 ICE facilities we examined, and considering all three scenarios, the cumulative number of ICU admissions by day 90 for over half of facilities would exceed hospital capacity within a 10-mile radius. The impact on hospital capacity is lower if the radius was expanded to 50 miles, hovering around 8% of facilities exceeding ICU demand. These models do not take into account other concerns that would strain ICE’s operational capacity at a time when staffing is likely to be a concern, such as the difficulty and risk to staff of repeatedly transporting and guarding detainees up to 50 miles distant from their facilities of origin. The timing and peak number of cases are driven by facility size, which also are important when considering preventive interventions. |
| Using a stochastic disease transmission model, we estimated the number of people infected, hospitalizations, and deaths across three transmission scenarios after a successful introduction of SARS-CoV-2 into the Kutupalong-Balukhali Expansion Site. The introduction of SARS-- CoV-2 into the Kutupalong-Balukhali Expansion Site or any other large refugee or internally displaced persons (IDPs) camp or settlement is likely to have serious consequences and overwhelm existing health systems. Even when transmission rates were assumed to be similar to that of influenza (low scenario), the necessary hospitalization capacities far exceeded the available capacities for the refugees in the expansion site in most simulations |

| |
|---|
| Using a WHO forecasting tool, this study has identified COVID-19 case load, according to severity, and potential health system needs in NW Syria, an area that has been subjected to severe levels of violence, displacement and health system disruption during the nine years of the Syrian conflict. The study models use three scenarios. The Camp-population Scenario describes a situation whereby unmitigated spread in crowded displacement camps will lead to total health system collapse within the first four weeks of an outbreak. Considering that such a scenario can occur concurrently with Scenario Two (or a worse All-population Scenario Three not presented here whereby doubling rates and clinical attack rates would be higher), the epidemic outcomes can be catastrophic |
| This article presents an exploration of the COVID-19 pandemic among migrants and asylum seekers in Mexico, based on the available epidemiological surveillance information. Despite the limitations of the data, it is possible to draw some general conclusions. The first noticeable result is that 0.27% of the suspect cases registered in the database corresponded to migrants originating from Central America, the Caribbean, Venezuela or an African country, a very high percentage if one considers that international immigrants in Mexico were only about 0.11% of the population in mid-2020 in Mexico. When it is taken into consideration that the majority of international immigrants in Mexico come from the United States (Giorguli-Saucedo et al., 2016), the difference is even more striking. The calculation of cases per 100,000 shows a similar picture, particularly in scenario 1. This scenario uses as denominator the estimated number of migrants who remained on the northern border of Mexico, waiting to start their asylum application procedures in the United States, in April 2020. This could be an underestimation of the actual number, as not all migrants aiming to cross are registered in the waiting lists that are the source of this data (Leutert et at., 2020)<br><br>In conclusion, according to our results, migrants and asylum seekers in Mexico are a group at risk for infectious respiratory diseases that, in the context of the COVID-19 pandemic, could be disproportionately affected |

| |
|---|
| The present study suggests that about three-fourth (73.5 %) of the participants screened positive for either depression or anxiety. All the migrants who screened positive for anxiety also screened positive for depression, suggesting high co-morbidity. Additionally, about one-fifth of the participants screened positive for only depression. Additionally, on the self-designed questionnaire, a significant proportion of participants reported a marked increase in negative emotions and feelings such as loneliness, tension, frustration, low mood, irritability, fear, fear of death, and social isolation |
| From early April 2020, a marked escalation of new cases of COVID-19 was observed among low-skilled migrant workers living in dormitories in Singapore. By 6 May 2020, these infected workers formed 87.9% of the 20 198 cases of cases of COVID-19 confirmed in Singapore.<br>Unsatisfactory housing and social overcrowding in accommodation for low-skilled migrant workers need to be addressed before any pandemic occurs. If this is not done, epicentres of the disease can arise in these housing areas. Once they occur, management of these localised outbreaks have to be swift and comprehensive, in order to avoid spillover infections to the general population |

To the best of our knowledge this is the first report of patients dying with TB and COVID-19, including 69 patients from the two largest cohorts of co-infected patients available so far. Although the case-fatality rate was rather high (overall 10.6%, but 14.3% in the first cohort) and still preliminary (it can increase over time within both cohorts), the results seem consistent with those observed in other cohorts of COVID-19 patients.1-3 In general, all patients (except one) were aged >65 years, and were affected by >2 comorbidities. In all cases COVID-19 contributed to worsen the prognosis of TB patients and/or to cause death.

In the present study, we observed that confirmation and hospitalizations were both more frequent in the states where more clinics and hospitals are available. At the same time, a proportion of patients moved from their state of residence to seek medical attention, according to the case-by-case results. Although they might seem discordant, our results show a lower confirmation and hospitalization in state-clusters with more availability of clinics and hospitals
From the beginning of the outbreak, migration was significantly associated with incidence ratios according to state-cluster analyses. Confirmation was not performed in all hospitalized patients, but 72.15% of hospitalized patients had positive outcomes to date. Elderly patients had lower odds of being hospitalized, but were likely to die, while interstate migrants had more propensity to fatal outcomes, yet were less likely to be laboratory-confirmed. Age group laboratory-testing, if not corrected, could result in biased assumptions of severity and lethality among young patients. These findings may help health professionals and policymakers to consider the importance of maintaining the shelter-in-place policy to prevent further exposure to communities not currently affecte

To our knowledge, this is the only study addressing SARS-CoV-2 carriage among different precarious populations including homeless adults but also children and other hard-to-reach populations during the COVID-19 outbreak in France. The strength of our study is its large population size, with a high (78.9%) acceptance rate toward testing, particularly among individuals living in precarious conditions (92.1%) suggesting that this population is concerned about the disease. We found an overall 7.0% SARS-CoV-2 positivity rate, with most infected individuals among homeless people and employees working in homeless facilities, while no cases were found in asylum-seekers and in other people also living in precarious conditions

The other major finding of our work is an apparent higher risk for COVID-19 for individuals from Sub-Saharan, Caribbean or Latin-American origin. All of them have equal access to the virtually universal health coverage available for Spaniards or migrants from other areas of the world.
In summary, we report a selective increased risk for COVID-19 among certain migrant populations in Spain: Sub-Saharan, Caribbean and Latin-America that is not related to unequal access to health care. These groups may deserve a particular attention, particularly when our country, as well as others, is beginning a de-escalation of social distancing measures

| |
|---|
| Whereas 21% of the cohort (of 5977 foreign workers) were found to be positive for COVID-19, the true prevalence is likely higher if asymptomatic individuals are also assessed, with incidence up to 36% in other closed environments.<br>The strategy for containment in closed-living environments therefore would be to isolate symptomatic individuals, and to establish public health measures for social distancing |
| In conclusion, the reactions presented here clearly show that Asian students in regions yet unaffected by SARS-CoV-2 could have already experienced anuncomfortable level of prejudice inthe public spaces encompassing transport, gastronomy, shopping, health services and university. Such behaviors can particularly affectthose individuals of Asian origin who are tending to wear face masks.<br>Overall, these findings underscore the responsibility of different parties in overcoming and preventing discrimination during outbreaks of infectious diseases. Firstly, universities that host Asian students and staff from abroad should support their students during the outbreak (and any other future epidemic emerging from Asian region) and protect them from harmful misconceptions. |
| This study revealed some specific socio-psychological experiences of respondents. However, it is also admirable that many of the international students were afraid during pandemic. This may be due to the fact that the respondents in affected areas paid more attention to the safety of their families [4]. Secondly, students with longer stay in China reported more concerns and consequences than the students who stayed for a short period of time. This may be associated with the respondent's age and their marital status |

| |
|---|
| Camp residents report COVID-19 symptoms almost twice as frequently as members of the host community. We also document differences in self-reported non- COVID-19 symptoms, but these are not statistically significant.<br>While this suggests that COVID-19 is much more prevalent in the refugee population,we cannot definitively exclude two alternative explanations. The first is that refugees experience higher rates of other common illnesses with overlapping symptoms. The second is that some refugees over report adverse life events and health outcomes, as some anecdotal evidence suggests. |
| Findings of this study suggest the following recommendations for future interventions: (1) more attention needs to be paid to vulnerable groups such as the young, the elderly, women and migrant workers; (2) accessibility to medical resources and the public health service system should be further strengthened and improved, particularly after reviewing the initial coping and management of the COVID-19 epidemic; (3) nationwide strategic planning and coordination for psychological first aid during major disasters, potentially delivered through telemedicine, should be established and (4) a comprehensive crisis prevention and intervention system including epidemiological monitoring, screening, referral and targeted intervention should be built to reduce psychological distress and prevent further mental health problems. |

| Generalisability/ External Validity (as reported) |
| --- |
| None explicitly stated. |
| - Awareness that not all refugees are based in camp-like settings and e.g. governmental measures to protect their citizens which exclude e.g. refugees, nonnationals |

| |
|---|
| None explicitly stated. |
| None explicitly stated. |

| |
|---|
| None explicitly stated. |
| None explicitly stated. |

| |
|---|
| The main limitation of this preliminary study is that the cohort, (…) cannot be considered representative either of the European nor of the global situation |
| None explicitly stated. |

| |
|---|
| None explicitly stated. |
| None explicitly stated. |

| |
|---|
| None explicitly stated. |
| None explicitly stated. |

| |
|---|
| This study is among the first documen ting the prevalence of COVID-19 symp toms and risk factors in a representative sample of both refuge and host communities. |
| - |

| Main limitations (as reported) |
|---|
| - A crucial limitation is our reliance upon estimates of the proportions of individuals requiring hospitalization and ICU access, across different ages, reported by a CDC report based on limited data: in time, these proportions may prove to be overestimates and the calculations we provide seen as overestimates<br>- The treatment capacity of local hospitals was calculated using data made public the first week of April, but actual capacity over time will vary depending on the rate at which the virus spreads in the general population, and we may therefore have underestimated need<br>- while data reporting the current number of ICE detainees may differ from those used in this analysis, they lack the facility-level specificity needed to calculate ICU bed demand.<br>- The actual age distribution of ICE detainees at each facility is unknown, and we relied on aggregate estimates from 2019<br>- We assumed a fixed contact rate over the course of the epidemic<br>- Our model included counties in their entirety if any part of them fell within the specified radii of 10 and 50 miles, so it is likely that community health care capacities within these radii are overestimated<br>- Overall, these assumptions suggest that our findings may have underestimated the number of infections and the number of facilities that will overwhelm the community ICU capacity.<br>- ingress/egress not considered<br>- model starts all outbreaks at the same time |
| - We are using a mass-action model, which tends to overestimate the size of outbreaks because populations are generally not closed and well mixed<br>- With the particular demographic characteristics and health status of refugee populations, like this one in Cox’s Bazar, we need to be cautious when developing guidance based on<br>previously estimated properties of SARS-CoV-2/COVID-19.<br>- |

| |
|---|
| - This forecasting report uses globally reported estimates for key parameters such as doubling rate, attack rates and case-fatality rates. It is possible for the situation in NW Syria to be different, either negatively or positively, thus substantially altering projections.<br>- Our study did not use age-specific projection (such as CFR).<br>- No population specific factors (age structure, prevalence of comorbidities...)<br>- The COVID-ESFT considers an exponential growth model, which does not reflect a real epidemic curve that is seen in epidemics and pandemics.<br>- only estimate for the first six (to eight) weeks of an outbreak from first case<br>- important impact factors (e.g. preparedness, expected future interventions) not taken into account<br>- |
| - Firstly, the variable we employed to identify migrants is only an approximation to our population of interest (mixed migrant flows aiming for the United States), so it is possible that some of those identified by it were already established in Mexico as documented immigrants or refugees. We tried to minimize this problem by restricting the analysis to states where mixed migrant flows concentrate, and to the nationalities that are more frequent among those seeking asylum in the United States, but some measurement error surely remained<br>- The same is true for the denominators used in calculating<br>the cumulative incidence, that were only an approximation to the real number of mixed migrant flow members |

| |
|---|
| - The present study was based on the use of brief screening instruments which although have low reliability and validity and the results needs to be interpreted<br>keeping this fact in mind.<br>- |
| - |

The main limitation of this preliminary study is that the cohort, although likely to report the vast majority of cases with TB and COVID-19 in the countries surveyed, cannot be considered representative either of the European nor of the global situation

- limitations that derive from the timing of the COVID-19-related outcomes
- demographic migration indicators may exclude illegal migratory movements
- health infrastructure (clinics and hospitals) reflects only availability of health resources but not actual access to those resources
- incidence and confirmation may underestimate the true incidence and confirmation due to underreporting and asymptomatic carriers.
- Lethality may be considered with caution, as it may vary from official registries, as authors employed both confirmed and suspected cases as the denominator
- we did not adjust our results according to comorbidity distribution in the studied population (but for: population age and gender composition)

| |
|---|
| - Our study population was not randomly and homogenously recruited.<br>- Participants’ medical histories and use of individual preventive measures were not documented.<br>- Individuals were not asked about anosmia and ageusia. |
| - First, we did not collect information regarding<br>mobility or social interactions that may well differ for different migrant populations.<br>- Second, we did not have local data regarding socioeconomic status, education or health conditions for migrants.<br>However, on aggregate, migrants from Romania, Ukraine or Morocco belong to a relatively lower socioeconomic stratum all along Spain and do not exhibit an increased risk for COVID-19 as opposed to migrants from Sub-Saharan Africa or Latin-America.<br>- There are limited data regarding different susceptibility to respiratory viruses from different ethnic backgrounds. Some studies have suggested a greater susceptibility of racial minorities to influenza. However, many studies have failed to dissociate clinical outcomes from socioeconomic issues, particularly unequal access to health care 6–9. Different susceptibilities to COVID-19 with a genetic background with unequal world distribution are just beginning to be explored |

| |
|---|
| - Nonetheless, this cohort is generally young (mean age: 33 years) and fit, with likely lower mortality risks as compared to residents in other closed-living environments such as nursing homes .<br>- We also acknowledge that a single negative test does not exclude COVID-19 infections. It may thus have to be assumed that all symptomatic individuals housed in the affected dormitory should be suspected cases. |
| None stated. |
| None stated. |

We also document differences in self-reported non-COVID-19 symptoms, but these are not statistically significant. While this suggests that COVID-19 is much more prevalent in the refugee population, we cannot definitively exclude two alternative explanations. The first is that refugees experience higher rates of other common illnesses with overlapping symptoms. The second is that some refugees over report adverse life events and health outcomes, as some anecdotal evidence suggests.

-

| Additional (main) limitations identified by review-team |
|---|
| - |
| - |

-

-

| |
|---|
| - |
| - |

| |
|---|
| - |
| - |

| |
|---|
| People were only tested once, through infected employees the virus could spread within the accommodation |
| - |

| |
|---|
| - |
| - |
| - |

| |
|---|
| - no confirmed COVID-19 cases measured, only self-reported data on symptoms likely to be COVID-19 |
| - |

| | Risk of Bias |
|---|---|
| **Funding sources/ conflicts of interest** | **Overall rating** |
| This study was funded by grants from the National Institute of General Medical sciences (P20GM125507) and the National Institute on Drug Abuse (2T32DA013911- 16A1)<br>The funding organizations had no role in the design and conduct of the study; in the collection, analysis, and interpretation of the data; and in the preparation, review, or approval of the manuscript.<br><br>Authors don't give a statement about conflicts of interest | LOW |
| PS and ST received some funding for this work from the Office of Foreign Disaster Assistance (OFDA), USAID (https://www.usaid.gov/who-weare/organization/bureaus/bureau-democracy-conflict-and-humanitarian-assistance/office-us, grant number 130492), and the Centers for Disease Control and Prevention (CDC; https://www.cdc.gov/, grant number 126280). The funders had no role in study design, data collection and analysis, decision to publish, or preparation of the manuscript<br><br>The authors have declared that no competing interests exist. | LOW |

| | |
|---|---|
| This study has not received funding<br><br>The authors have declared no competing interest. | HIGH |
| Conflict of interest: none.<br>Funding sources: none | HIGH |

| | |
|---|---|
| We have no financial disclosure to make.<br><br>The authors declare that they have no conflict of interest | HIGH |
| The authors have not declared a specific grant for this research from any funding agency in the public, commercial or not-for-profit sectors.<br><br>Competing interests: None declared | LOw |

| | |
|---|---|
| This research did not receive any specific grant from funding agencies in the public, commercial, or not-for-profit sectors.<br><br>The authors have no conflicts of interest to declare. | HIGH |
| This research received no external funding.<br><br>The authors declare no conflict of interest. | MODERATE |

| | |
|---|---|
| This work had received financial support from the Fondation Méditerranée Infection.<br><br>No potential conflict of interest relevant to this letter was reported | HIGH |
| No funding was available for this manuscript.<br><br>The authors have declared no competing interest. | MODERATE |

| | |
|---|---|
| Conflict of interest statement None declared.<br>Funding sources None. | LOW |
| This research received no external funding<br>The authors declare no conflict of interest. | HIGH |
| Not stated. | HIGH |

| | |
|---|---|
| **Funding**: This study was funded by the Yale Y-RISE initiative, the Yale Macmillan Center's Program on Refugees, Forced Displacement, and Humanitarian Responses, and Innovations for Poverty Action's Peace and Recovery Initiative.<br>- **Conflict of Interest**: None declared | LOW |
| **Funding** The authors have not declared a specific grant for this research from any funding agency in the public, commercial or not-for- profit sectors.<br><br>**Competing interests** None declared. | MODERATE |
