## APPENDIX_C1 for "SARS-CoV-2 among migrants and forcibly displaced populations: a rapid systematic review": APPENDIX_C1_RoB_observ_studies.pdf

**APPENDIX C1:**
**Risk of Bias assessement of observati**

| Study Id | Reviewer | SELCTION Judgement |
|---|---|---|
| Motta et al. 2020 | MH/KB | HIGH |
| Mendez-Dominguez et al. 2020 | MH/KB | LOW |
| Koh 2020 | MH/KB | LOW |
| Chew et al. 2020 | HG/MH | LOW |
| Guijarro et al. 2020 | MH/KB | HIGH |

| | | |
|---|---|---|
| Ly et al. 2020 | MH/KB | HIGH |
| Bojorquez et al. 2020 | MH/KB | HIGH |
| Rzymski et al. 2020 | MH/KB | HIGH |
| Fakhar-e-AlamKulyar et al. 2020 | MH/KB | HIGH |

| | | |
|---|---|---|
| Kumar et al. 2020 | MH/KB | HIGH |

**Studies from snowball search**

| | | |
|---|---|---|
| Lopez-Pena et al. 2020 | MH/KB | LOW |
| Qiu et al. 2020 | MH/KB | UNCLEAR |

### onal studies using the EPHPP-tool

Quotes

*Data of both cohorts were combined to assess the mortality. Cohort A included 49 patients with TB and COVID-19 from 26 centres in Belgium, Brazil, France, Italy, Russia, Singapore, Spain, and Switzerland, 7 whereas cohort B included 20 cases admitted to a single reference hospital located in Northern Italy. The main limitation of this preliminary study is that the cohort, although likely to report the vast majority of cases with TB and COVID-19 in the countries surveyed, cannot be considered representative either of the European nor of the global situation.*

-

*Information was gathered from daily reports provided by the Ministry of Health, Singapore. 3 These reports provided information on the number of newly diagnosed cases, and the distribution of the cases among imported cases who arrived to Singapore from overseas, and non- imported cases. The non-imported cases are categorised into Community Cases (residents and employment pass holders excluding work permit holders and dormitory residents), work permit holders not residing in dormitories and work permit holders residing in dormitories.*

*on April 5 th [2]. We describe results of the outbreak investigation and clinical evaluation in dormitory A. This study is a review of investigation results from April 11 th to April 19 th, 2020. Residents in dormitory A who had symptoms of acute respiratory infection were assessed for COVID-19.*

*In all, 1832 out of 5977 foreign*

*Case was defined as a patient with a COVID-19 diagnosis at Hospital Universitario Fundación Alcorcón confirmed by Polymerase Chain Reaction (PCR) for SARS-CoV2 for adults residing in Alcorcón. Incident cases of COVID-19 were obtained from the Electronic Patient Record (Selene ©) that is used for all medical and administrative interactions with patients. COVID-19 clinical diagnosis was established by the attending clinicians at the Emergency Room according to European Center for Disease Control and Prevention (ECDCP)-World Health Organization Criteria (WHO)*
*The population at risk was defined as all adults included in the official municipal*
*live registry of population of the City Council of Alcorcón (last updated March 14, 2020)·*
*Incident cases of COVID-19 were obtained from the Electronic Patient Record (Selene ©) that is used for all medical and administrative interactions with patients.*

*four shelters (A-D) and four hotels (1-4), other people living in precarious condition (housed in residences α and β ), asylum-seekers (housed in residence γ ), and employees working in these accommodation centres.*

*Residence γ is dedicated to asylum-seekers, including family groups and single individuals.*

*Participants were encouraged by the management staff of the facilities to get tested and were then recruited on a voluntary basis.*

*Monitoring Units (USMER), throughout the country. USMER are first, second and third level care units from the different public health systems in Mexico, which report suspect cases of respiratory disease for which samples for laboratory testing are collected. Following the surveillance protocol, these units take samples from 10% of ambulatory cases and 100% of hospitalized patients that meet the COVID-19 suspect case definition. Non-USMER units take samples of severe suspect cases of COVID-19, which are also recorded in the database*
*information on the downloading site,these are preliminary data (not yet validated) collected by the Epidemiological Surveillance System for Viral Respiratory Disease (previously Epidemiological Surveillance System for Influenza -SISVEFLU-), which are captured by 475 Viral Respiratory Disease Monitoring Units (USMER), throughout thecountry. USMER are first, second*
*asylum in the United States.The data base from the Ministry of Health includes a dichotomous variable coded migrant/non- migrant. This variable does not indicate migration status (regular vs. irregular), and the information is missing in 99.5% of cases. Therefore, to identify our population of interest, we used the variable "country of nationality", and selected those whose nationality corresponded to one of the main sending countries or regions of mixed migrant flows in Mexico: Central America, the Caribbean, Venezuela and Africa (Cobo & Fuerte, 2012; Rodriguez, 2016). Furthermore, we restricted the analysis to suspect cases whose place of residence as registered in the database corresponded to any of the five states of the northern border region (Baja California, Sonora, Chihuahua, Coahuila and Tamaulipas) and the southern border region (Chiapas) where mixed migrant flows are concentrated. The rationale for this restriction was the assumption that foreign nationals of the aforementioned regions and countries staying in these six states would be more likely to belong to the population of interest, than foreign nationals in other states of the country.*

*The selected group included Asian students at Poznan University of Medical Sciences in Poland who were directly invited to complete the survey via an e-mail message. All surveyed students were living in the city of Pozna ´ n (Greater Poland Voivodeship).*

*The survey was completed by eighty-five medical students from Asia (mean ± SD age 23.8 ± 3.8; 49 females, 36 males), mostly from Taiwan (75.3%), who study at Poznan University of Medical Sciences in Poland. All of them have been living in Pozna ´ n for at least half a year (with a mean ± SD of 2.7 ± 1.4 years).*

*We approached students through official WeChat groups, which were already developed by the universities for international students.*

*sent was obtained before recruitment. To be included in the study, the participants were required to be aged > 18 years, of any gender, able to understand Hindi, were co-operative, medically stable, and provided the written informed consent*

*It was a cross-sectional study conducted in the Chandigarh, a Union Territory, in North India. The migrants' workers identified by the Government of India, who were living in the shelter house or government authorized buildings, were recruited.*

---

*We administered a phone-based survey to a sample of 1,255 households between April 11 and 17, 2020 to assess the health status, health behaviors, and livelihoods of households across Cox's Bazar. Of those, 909 were reachable by phone and 899 consented to be surveyed. The sample for this study was taken from the Cox's Bazar Panel Survey (CBPS), a longitudinal study tracking 5,020 households across Cox's Bazar that is divided almost equally between refugee camps (n=2,493) and host communities (n=2,527). The Primary Sampling Units (PSUs) in host communities are mauzas, the lowest administrative unit in Bangladesh.*

*This study is the first nationwide largescale survey of psychological distress in the general population of China during the tumultuous time of the COVID-19 epidemic. [...] Leveraging the Siuvo Intelligent Psychological Assessment Platform, we presented QR codes of the questionnaire online openly accessible to the general public nationwide*

---

| Comments | STUDY<br>Judgement | Quotes |
|---|---|---|
| No sampling, no denominators, unclear who / what the target population is, selective sample from a Network | HIGH | - |
| Population based data from official statistics | HIGH | - |
| National notification data used, and target population is well described and registered | LOW | - |
| The selected individuals are described as a cohort of foreign Workers (n=5977) living in a dormitory residence in Singapore; there is no further definition of "foreign workers", yet this description is suffienct to assume that the selected individuals are representative of our target population (i.e. labor migrants). As this is a cohort analytic study, there is no participation rate. | LOW | - |
| No sampling, registry data with self-referrals to one single hospital. | HIGH | - |

| | | |
|---|---|---|
| Unclear how many shelters there are and how those studies were selected, including enrolled invidiuals. | HIGH | *A cross-sectional survey* |
| National coverage, surveillance sites, first, 2nd and 3rd level facilities (not only hospitals) - still not population based, but acceptable, because primary care sites seem to be included. However, irregular migrants are identified by a proxy variable of nationality and place of residence in the border region - this is very much prone to misclassification bias for the initial population of interest. | LOW | - |
| No denominators reported of target population at Poznan University vs overall Asian Students in Poland, no random sampling procedure | HIGH | *The survey data collected anonymously do not require approval by the local bioethical committee in Poland. Potential respondents were informed in the invitation message about the general subject matter of the survey, its voluntary and fully anonymous charac- ter (including no IP tracking)* |
| Non-random sample, no mention of denominators | HIGH | *purpose, a cross sectional study* |

| | | |
|---|---|---|
| No random sampling reported, no denominators reported | HIGH | - |
| Cantacted people from the Cox’s Bazar Panel Survey (CBPS), should be quite "representative". | MODERATE | - |
| As a nationwide survey we assume data to be "representative". The Question, if it is "representative" for the migrant worker population remains. | MODERATE | *This study is the first nationwide largescale survey of psychological distress in the general population of China during the tumultuous time of the COVID-19 epidemic. A self-report questionnaire was designed to survey peritraumatic psychological distress during the epidemic.* |

| Comments | CONFOUNDE<br>Judgement | Quotes |
|---|---|---|
| No follow-up time reported, cohort design | UNCLEAR | *17/20 (85.0%) in cohort B. <b>Migrants were younger than natives: in cohort A the median (IQR) age was 40 (27- - -49) VS. 66 (46- - -70) years, whereas in cohort B 37 (27- - -46) VS. 48 (47- - -60) years. Overall, migrants had a lower mortality rate than natives (1/43, 2.3% versus 7/26, 26.9%; p-value: 0.002). Migrants had fewer co-morbidities than natives; in par- ticular, 23/43 (53.5%) migrants had no co-morbidities versus 5/26 (19.2%) natives (p-value: 0.005).</b> Among the patients who died,* |
| Aggregate, population-based data, ecological design, no spatial analysis | HIGH | - |
| National notification data, analysed in a cross-sectional design over a given period, due to national scope | HIGH | - |
| Cohort analytic study: cohort of foreign workers residing in a dormitory (n=5977), there is a comparison Group (covid-19-positive vs. covid-19-negativ workers are compared for different Symptoms). | HIGH | - |
| Cross-sectional study using single-centre hospital data with the aim to assess different incidienc risks depending on region/country of origin among residents in Alcorcon, so migration is exposure. Rather weak study design | LOW | *Multivariate negative binomial regression model with robust variance was used to estimate the incidence rate by world zone adjusted for sex and age.* |

| | | |
|---|---|---|
| One-off, cross-sectional testing / screening of shelter inhabitatnts | HIGH | *factors*<br><br>*people*<br><br>*A separate multivariate logistical regression analysis was used to identify independent risk factors for SARS-CoV-2 carriage prevalence among all individuals and in selected groups (when positive cases were found).* |
| Study design judged in itself not against RCT standards, for observational study it is a robust design | LOW | *The model was adjusted for sex, age and the presence of at least one risk condition (hypertension, diabetes, chronic obstructive pulmonary disease-COPD, asthma, obesity or pregnancy). Missing data were handled with pairwise deletion. Lastly, the cumulative incidence in* |
| Online cross-sectional study with self-selected individuals | HIGH | - |
| Cross-sectional study | HIGH | - |

| | | |
|---|---|---|
| Cross-sectional survey in one region in North India, unclear sampling strategy, no random sample | HIGH | *Further, the findings are twice that seen in an online survey of the general population, done during this lockdown period (Grover et al., 2020). If we compare to the findings of the online survey done during the lockdown period, another important fact, which is evident from this study, is that a higher proportion of the participants screened positive for depression, rather than the anxiety. These differences possibly suggest different psychological reactions of people belonging to dif- ferent socioeconomic strata. The online survey perhaps included people of middle and higher income who probably had a higher level of an- xiety, that could be related to the ongoing pandemic per se.* |

| | | |
|---|---|---|
| Comparison between host community and refugee households given, no randomization mentioned. | LOW | *[…] after adjusting for basic sociodemographic characteristics and pre-COVID living conditions, such as toilet sharing, employment, and household assets.* |
| - | HIGH | *Multinomial logistic regression analyses showed that one's CPDI score was associated with their gender, age, education, occupation and region.* |

| Comments | **DATA** Judgement |
|---|---|
| Descriptive analysis, not really applicable | HIGH |
| Strata at aggregate level considered, but overall residual confounding still seems possible | LOW |
| Overall, stratified reporting by type of housing / dormitoris vs. nationals, but no further stratification was done, so there migt be some confounding in principle, however, it is not sure, if any factor would affect the differences to an extent that diminshes them | LOW |
| No confounders considered (socio-demographics, context, work environment, individual behaviour, etc) | LOW |
| Adjusted for age and sex, but severity of disease has not ben controlled for, which may lead to less utilisation e.g. by Spaniards in case e.g. Afrcian migrants have more sever course of disease they will be overrepresented in the data | LOW |

| | |
|---|---|
| See table 4, migration was only analysed in unvariate analysis, unclear why, but risk of residual confounding therefore high | LOW |
| Adjustment for major co-variables | HIGH |
| Descriptive reporting, no stratification by age, sex as minimum confounders, some other issues considered (wearing mask vs not etc) | HIGH |
| No counfounders reported, unclear, if taken into consideratoion. | HIGH |

| | |
|---|---|
| No stratification by age, sex, SES, caste, etc | HIGH |

| | |
|---|---|
| Adjusted for confounders | LOW |
| Adjustment for confounders unclear therefore we assessed high risk of bias. | MODERATE |

| Quotes | Comments |
| --- | --- |
| - | Retrospective data collection, clinical records, self-reports |
| - | Population based data from official statistics,PCR-tests as valid instrument to test for SARS-CoV-2 |
| *In Singapore, cases of COVID-19 are confirmed by a posi- tive finding of a nasal swab which undergoes PCR testing for SARS- CoV-2.* | PCR-tests as valid instrument to test for SARS-CoV-2 |
| *Symptoms and clinical signs were collected, with fever defined as 37.6 C. Oxygen saturations and heart rate were measured using portable pulse oximeters. Nasopharyngeal specimens were sent for polymerase chain reaction (PCR) testing for severe acute respiratory syndrome coronavirus-2.* | PCR-tests as valid instrument to test for SARS-CoV-2 |
| *as a patient with a COVID-19 diagnosis at Hospital Universitario Fundación Alcorcón confirmed by Polymerase Chain Reaction (PCR) for SARS-CoV2 for adults residing in Alcorcón.*<br>*For molecular diagnosis of SARS-CoV-2 infection, nasopharyngeal swabs, sputum, or bronchopulmonary aspirates were processed by automatized extraction using the MagNaPureLc instrument (Roche Applied Science, Mannheim, Germany) and real time reverse-transcription PCR using the SARS-Cov-2 nucleic acid detection Viasure kit (CerTestBiotec S·L·), following the manufacturer's instructions. For this rRT-PCR, we used Bio-Rad CFX96™ Real-Time PCR Detection System. We amplified two different viral regions: ORF1ab gene (FAM channel), N gene (ROX channel), and the internal (HEX channel). Cycle threshold values ≤ 4 0 were considered positive. Positive and negative controls were included in each run for every assay.* | PCR-tests as valid instrument to test for SARS-CoV-2 |

| | |
|---|---|
| *They were systematically asked to provide basic demographic information (sex, age, country of origin), chronic conditions, and any respiratory symptoms or fever in the two weeks prior to sampling. Body temperature was measured using a forehead infrared thermometer. Nasal samples were systematically collected on transport media using Sigma Transwabs (Medical Wire, Corsham, United Kingdom). For self-sampling, participants were invited to insert the swab into their nostrils (about 2 cm). If individuals were unable to perform self- sampling, trained investigators carried out the sampling. Specimens were immediately processed for SARS-CoV-2 PCR testing. Homeless peoples' pets were also tested with the approval of their owner and their nasal swabs were collected by vets.*<br>*Real-time reverse transcription-PCR amplification was used to confirm the presence of SARS- CoV-2 RNA targeting the gene coding for the envelope (E) protein, as previously described* | PCR-tests as valid instrument to test for SARS-CoV-2, but symptoms, and migration and asylum status unclear how it was measured |
| *be part of this subpopulation. Thus, the cumulative incidence among migrants was calculated as the number of suspect cases in this group in the five states on the northern border, divided by the number of people estimated to be on the northern border waiting for their migratory procedures to begin or be resolved. This last estimate was taken from two different sources, from which two scenarios were constructed. Scenario 1 corresponds to the suspect cases, divided by the 14,400 migrants who, according to a study by the University of Texas at Austin and the University of California at San Diego, were waiting to submit their asylum application in April 2020 as a result of the metering system (Leutert et al., 2020). Scenario 2 took as denominator the number of cases in the United States immigration courts assigned to the MPP program from March 2019 to March 2020, which corresponded to people of the nationalities defined for our work (59,346), according to a project by Syracuse University (Syracuse University, 2020). For people of Mexican nationality, the population at mid-2020 minus the number of international immigrants was used as the denominator, with data from the National Council of Population in Mexico (CONAPO) available at http://www.conapo.gob.mx/work/models/CONAPO/Mapa_Ind_Dem18/index.html.*<br><br>*or the metering system stay at the border sometimes for months, so that those registered as suspect cases of COVID-19 are more likely to be part of this subpopulation.* | Many proxy measures, assumptions, and constructed variables for migation status |
| *a self-designed, structured questionnaire was conducted* | No description of tool development, pretesting, validation etc |
| *was developed by using a questionnaire.*<br>*Sociodemographic characteristics of the respondents. b; COVID-19 impact on student life (N; number of total respondents, n; refers to respondents who answered "Yes" to the question). c; Psychosocial experiences of participants during the COVID-19 outbreak (1; Frequency of talk about COVID-19, 2; Fear of COVID- 19, 3; Worried about family members, 4; Depression about the pandemic, 5; Feeling Helplessness). d; Depicts COVID-19-prevention measures took by the inter- national students (1; Postponed visit to affected areas, 2; Decreased contact with others, 3; Increased care of washing hands, 4; Increased care of room ventilation, 5; Decreased visits to public gathering).* | Self-developed tool |

| | |
|---|---|
| *We used two brief screening instruments i.e. Patient Health Questionnaire-2 (PHQ-2) (Kroenke et al., 2003) and Generalized An- xiety Disorder- 2 (GAD-2) (Skapinakis, 2007) to assess depression and anxiety respectively. Both these scales have been used previously in many studies with adequate sensitivity to screen depression and anxiety (Hughes et al., 2018; Whooley et al., 1997).Perceived stress scale-4 (PSS-4) was used to assess perceived stress which has been reported to be the most useful and feasible in the situations where a short ques- tionnaire is required such as telephonic interview (Lee, 2012). Ad- ditionally, a self-designed questionnaire was used to assess the emo- tional and behavioural response to the lockdown. All the participants were administered these questionnaires by a trained Clinical Psychologist.* | Used both: established and self-developed tools. No reporting how the other tools were developed, pretested, etc. |
| *We administered a checklist of symptoms based on the WHO and CDC guidelines. We used the three most common symptoms featured on the WHO dedicated COVID-19 website on April 27, 2020 to produce our preferred measure of COVID-19 risk: having at least one of the symptoms (fever, dry cough, and fatigue or tiredness)* | Valid an reliable instruments were used, only the list of symptoms is a bit vague. The method of phone based interviews leaves potential to selection or social desirability bias |
| *We measured lifetime trauma using the Harvard Trauma Questionnaire (HTQ)8 and depressive mood using the 9-item version of the Patient Health Questionnaire (PHQ-9). We used a cut-off point of PHQ-9 equal to or above 10 as a screener for depression.* | Valided instruments were used, yet method of online questionnaire raises concerns to risk of selection bias. |

| WITHDRAWALS AND DROP-OUTS | | | OVERALL | |
|---|---|---|---|---|
| Judgement | Quotes | Comments | Judgement | Comments |
| HIGH | - | not reported | HIGH | Overall high risk of bias in almost all domains |
| UNCLEAR | - | not applicable | MODERATE | Low risk of bias in Data collection methods and Selection bias, but high risk in study design and confounders, overall a moderate risk of bias |
| UNCLEAR | - | not applicable | LOW | Overall low risk of bias in almost all domains only a high risk with confounding factors |
| UNCLEAR | - | not applicable | LOW | Overall low risk of Bias only confounding factors are not considered |
| UNCLEAR | - | not applicable | MODERATE | Study design and Selection of population indicate a high risk of bias whereas Data collection methods and confounding factors point out low risk of bias. Overall we rate a moderate risk of bias for this study |

| | | | | |
|---|---|---|---|---|
| UNCLEAR | - | not applicable | HIGH | Almost all domains indicate a high risk of bias. Data collection methods seem to be at low risk of bias, even though some methods remain unclear |
| UNCLEAR | - | not applicable | HIGH | We identified selection of study population and data collection methods at high risk of bias, confounders and study design at rather low risk of bias. We rate a overall high risk of bias, because the items that are the basis for data analysis are at high risk of bias. |
| UNCLEAR | - | not applicable | HIGH | Overall high risk of bias in all domains |
| UNCLEAR | - | not applicable | HIGH | Overall high risk of bias in all domains |

| | | | | |
|---|---|---|---|---|
| UNCLEAR | - | not applicable | HIGH | Overall high risk of bias in all domains |

| | | | | |
|---|---|---|---|---|
| UNCLEAR | - | not applicable | LOW | Well designed population-based cross-sectional study, overall risk of bias is low, though study design is cross-sectional so scores low on EPPH |
| UNCLEAR | - | not applicable | MODERATE | Difficult to appraise as quality of reporting is low. |
