## APPENDIX_C2 for "SARS-CoV-2 among migrants and forcibly displaced populations: a rapid systematic review": APPENDIX_C2_RoB_model_studies.pdf

### APPENDIX C2
### Risk of Bias Assessment for modelling studies

| Study information | | | Model structure | |
|---|---|---|---|---|
| Reviewer initials | Checked by | Study ID | 1. Are the structural assumptions transparent and justified? | 1. Comments |
| Overall | MH/KB | Irvine 2020 | No to minor concerns | transparent and well jsutified |
| Overall | MH/KB | Trulove 2020 | No to minor concerns | transparent, but partly they seem very pragmatic |
| Overall | MH/KB | Hariri 2020 | Moderate concerns | partly, the fundamental approach of the model remains unclear |

| 2. Quotes | 2. Are the structural assumptions reasonable given the overall objective, perspective and scope of the model? |
|---|---|
| *We implemented a simple stochastic susceptibleexposed-infected-recovered (SEIR) model. In this model, individuals are initially susceptible (S) to COVID-19 infection and become infected and transition to an exposed class (E) at a rate dependent on the reproductive number, known as the R0, and the proportion of individuals who are currently infectious (I). Individuals in the exposed class then transition to an infectious state (I) after a latent period and, finally, to a recovered state (R) at a constant recovery rate. The SEIR model and related variants support the vast majority of COVID-19 modeling studies around the world at present.* | No to minor concerns |
| *We used a stochastic Susceptible Exposed Infectious Recovered (SEIR) mathematical model to simulate transmission in this population [15]. To capture the potential variability of transmission possible in this setting, we simulated epidemics under three potential scenarios with different values of the basic reproductive number, R0:* | No to minor concerns |
| - | No to minor concerns |

| | Input data | |
|---|---|---|
| 2. Comments | 2. Quotes | 3. Are the input parameters transparent and justified? |
| assumptions are fitting to current state of knowlegde | - | No to minor concerns |
| assumptions are fitting to current state of knowlegde | - | No to minor concerns |
| different scenarios for different populations and transmissions respectively | - | No to minor concerns |

| 3. Comments | 3.Quotes | 4. Are the input parameters reasonable? |
|---|---|---|
| - | *Drawing from the published estimates, we studied three scenarios ranging from a most optimistic to most pessimistic estimate of the R0 as 2.5, 3.5, and 7 for low, medium, and high R0 scenarios, respectively. The incubation and infectious periods were estimated to be 6.4 days [16] and 7 days [17], respectively.* | Moderate concerns |
| yes, see table 1 | - | Moderate concerns |
| reported in table 1 | - | Moderate concerns |

| | Validation (external) |
|---|---|
| 4. Comments | 5. Has an external validation process been described? |
| Parameters may not be 1:1 transferable, but the validity of the scenario in terms of ist impact (what if this parameter would be present in detention centres) remains unaffected | Moderate concerns |
| Data is taken primarily from Wuhan outbreaks, which raises questions on transferability to crowded camp settings | Moderate concerns |
| Heterogeniety of populations not considered | Major concerns |

| | | | Validation (internal) |
|---|---|---|---|
| 5. Comments | 6. Has the model been shown to be externally valid? | 6. Comments | 7. Has an internal validation process been described? |
| No process, most data relies on "real" data, but no external validation per se, especially related to covid data in detention | Major concerns | not reported | Major concerns |
| In author contributions "Validation" is listed, although ist unsure where ist reffered to. We rate rather moderate than major concerns. | Major concerns | not reported | Moderate concerns |
| not reported | Major concerns | not reported | Major concerns |

| 7. Comments | 8. Has the model been shown to be internally valid? | 8. Comments |
|---|---|---|
| not reported | Major concerns | not reported |
| In author contributions "Validation" is listed, although ist unsure where ist reffered to. We rate rather moderate than major concerns. | Major concerns | not reported |
| no information /not reported | Major concerns | no information /not reported |

| Uncertainty | | Transparency |
|---|---|---|
| 9. Was there an adequate assessment of the effects of uncertainty? | 9. Comments | 10. Was technical documentation, in sufficient detail to allow (potentially) for replication, made available openly or under agreements that protect intellectual property? |
| Moderate concerns | Three scnearios, a probabilistic linear approach would have been stronger | No to minor concerns |
| Moderate concerns | 3 scenarios, a more probabilistic scenario would have been stronger | No to minor concerns |
| Major concerns | No confidence intervals for estimates, rigid approach with three scenarios | Major concerns |

| | OVERALL Risk of Bias: | |
|---|---|---|
| 10. Comments | Judgement | Comment |
| Minor concerns as very detailed in the text which allows for replication | LOW | Overall, this study has a low risk of bias, although validation raises concerns, the model is well reported and Input data is appropriate |
| supplement provided and details in methods | LOW | Overall, this study has a low risk of bias, although validation raises concerns and reasonability of input data might be discussed. |
| Overall documentation, level of details and methods are not sufficient to allow replicability | HIGH | Overall, high risk of bias for major and moderate concerns exist in 8 out of 10 domains. Moreover the replicability of the model as well as the fundamental approach to the model is not sufficently reported. |
